# CLUSTERING OF RARE VARIANTS FOR CAUSAL VARIANTS IDENTIFICATION AND EFFECT DIRECTION CLASSIFICATION

**DOI:** 10.1101/2024.02.22.24303151

**Authors:** Xianbang Sun, Xue Liu, Chunyu Liu

## Abstract

Several gene-based tests, e.g., sequence kernel association test, have been developed for association testing of rare single nucleotide variants (SNVs) in genomic regions with disease traits. A common limitation of these aggregate methods is their inability to discriminate potentially causal variants from null variants within the tested regions. We propose a novel clustering method to classify rare variants into null and signal variant groups using summary statistics from the gene-based tests based on a Gaussian mixture model (GMM). We classify the signal variants into potentially risk and protective subgroups of different effect sizes. We evaluate the performance of the proposed method by a simulation study, considering several statistics such as the adjusted rand index (ARI), mean square error (MSE), and accuracy in specifying the number of clusters. We apply the proposed clustering method to identify possibly risk and protective rare variants in six genes that are significantly associated with blood pressure (BP) traits in the most recent large genomewide association study (GWAS) and meta-analysis. This proposed method may facilitate the identification of potentially causal rare variant clusters in genomic regions and ultimately help understand the genetic architecture underlying human complex traits for the discovery of drug target and the design of gene therapy.

## INTRODUCTION

During the last decade, genome-wide association studies (GWASs) have identified hundreds of thousands of common genetic variants (minor allele frequency ≥ 5%) associated with numerous complex diseases and quantitative traits.^1,2^ in addition, low-frequency variants (1% ≤ MAF < 5%) and rare variants (MAF < 1%) are detected increasingly with the advent of next-generation sequencing technologies. Low-frequency variants and rare variants are substantial sources of unexplained heritability of various phenotypes.^3^ Several gene-based tests such as the burden test^4^ and sequence kernel association test (SKAT)^5^ have been developed for association testing of rare single nucleotide variants (SNVs) in genomic regions with disease traits. It has been shown that none of these gene-based tests is uniformly most powerful. Hence, omnibus tests such as SKAT-O^6^ and aggregated Cauchy association test (ACAT-O)^7^ are proposed to search for an optimal combination of the burden test and SKAT to provide robust summary statistics. Unlike single/multiple variant models, a common limitation of these aggregate methods is that they do not discriminate potential causal variants from null variants in association testing within the tested regions.

To overcome this weakness, we propose a clustering method based on a Gaussian mixture model (GMM) to discriminate potentially causal rare variants from null variants in the gene regions that are associated with disease traits. In large GWAS and meta-analysis, gene-based association analyses (e.g., SKAT-O and ACAT-O) are conducted to combine the burden test and SKAT to detect significant genes or regions (deemed as “signal regions” in this dissertation) in which rare variants show associations with a trait. The burden test outperforms SKAT when a large proportion of rare variants within a region are trait-associated, and most of the trait-associated rare variants have the same effect direction. The SKAT has a larger power than the burden test otherwise.^5^ For a given signal region associated with a disease trait, we fit multiple-variant models to obtain association statistics between phenotype and rare variants within the region. Based on the variant-level statistics, this novel clustering method may identify potentially risk and/or protective genetic variants in a genomic region. Furthermore, the method may also cluster the signal variants into subgroups of variants with different effect sizes and effect directions in association testing. We simulate genomic regions with independent rare variants and variants in linkage disequilibrium (LD). We evaluate the performance of the proposed method by a comprehensive simulation study with several statistics, including the adjusted rand index (ARI), mean square error (MSE), and accuracy of the number of clusters specification. We then apply the new clustering method to identify risk and protective rare variants in six genes that are significantly associated with blood pressure (BP) traits in the most recent large GWAS and meta-analysis.^8^ The identification of risk and protective rare variant clusters is critical not only for investigating the underlying biological mechanism between rare variants and disease traits, but also for the identification of drug targets and the design of gene therapy.

## METHODS

### Association testing of rare variants

The burden test^4^ collapses the rare variants within a gene region into a single genetic score and then tests the association of that variable with the trait. SKAT^5^ employs a score-type variance component test. The test statistics of these two tests can be constructed by the score statistics of rare variants in the tested region. The score statistics of j^th^ rare variant are given by

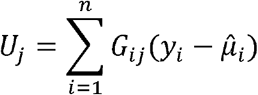

Where *G*_*ij*_ is the genotype or genetic dosage of the i^th^ individual at the j^th^ locus,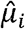 is the estimated mean of the outcome *y*_*i*_ under the null hypothesis that there is no association between the variants in the gene region and the phenotype. The test statistic of the burden test is 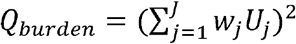, where *J* is the number of rare variants within the target region and *w*_*i*_ is the weight of the j^th^ variant. Under the null, *Q*_*burden*_ follows an asymptotic chi-square distribution with 1 degree of freedom.

The burden test is most powerful when all rare variants in the target region have the same effect direction and similar effect sizes. The burden test is underpowered when a large proportion of rare variants are non-causal or have opposite effect direction. The SKAT outperforms the burden under such scenarios. The SKAT test statistic is 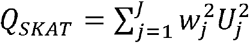 which follows a mixture of independent chi-square distributions asymptotically with 1 degree of freedom under the null. To improve power by combining these two tests, we adopt the ACAT-O method^7^ with two commonly used choices of weights based on MAF of beta density. The test statistic of the ACAT is 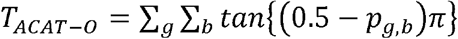, where *g* ∈ {*burden,SKAT*} *b* ∈ (*beta* (1,1),*beta* (1,25)}.The p-value of ACAT is calculated by 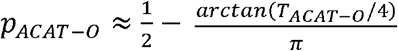.The clustering analysis of rare variants is performed only if the p-value of ACAT-O is smaller than a selected threshold, i.e., *p*_*ACAT* −0_ ≤ *α*, for a given region.

### Multiple-variant model to obtain variant level summary statistics

If a gene region shows statistical significance with a disease trait (*p*_*ACAT* −0_ ≤ *α*), we perform multiple-variant analysis for all rare variants within the region to account for potential LD between the rare variants. By adjusting for covariates, the multiple-variant model is given by

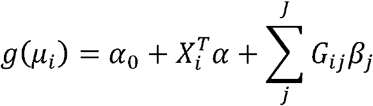

where *g* (·)is the identity function for a continuous trait and the logistic function for a binary trait.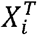 is a row vector of covariates of the i^th^ individual, *G*_*ij*_ is the genotype of this individual at the j^th^ locus within the target gene region. The beta coefficient 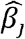, standard error 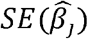 and corresponding p-value *p*_*j*_ of the j^th^ rare variant are obtained from the multiple-variant model. The beta coefficient and standard error statistics are used for the subsequent clustering analysis.

### Gaussian mixture model

The Gaussian mixture model assumes data is generated from a mixture of Gaussian (normal) distributions. We assume that each beta coefficient from a multiple-variant model has its variance which is estimated by 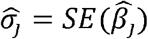. We also assume that each beta coefficient can be classified into two types of clusters: a null cluster and K signal clusters. Therefore, the total number of clusters is K^*^=K+1. A null cluster includes all non-causal rare variants of a gene region. Within each of the K signal clusters, the causal variants have a similar effect on the trait. With a null cluster and K signal clusters, each beta coefficient *β*_*j*_ of the multiple-variant model follows one of the normal distributions of 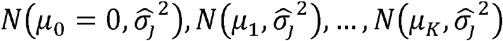, where *μ*_*k*_ is the true mean of beta estimates for rare variants classified in the same cluster.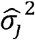 is the estimated variance of the j^th^ beta coefficient from the multiple-variant model, *μ*_0_ =0 is the mean of the null cluster. For each*β*_*j*_, we introduce a hidden (unobserved) class label variable *D*_*j*_ which follow a categorical distribution (that is the generalized Bernoulli distribution). The class label variable *D*_*j*_ indicates the component (cluster) that 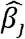 belongs to; that is,

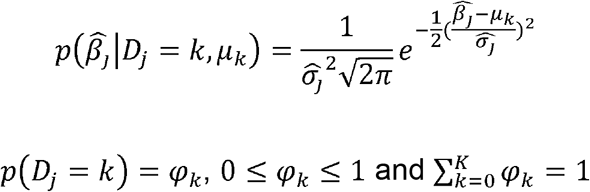

*φ*_*k*_ is the proportion of component *k* in the mixture distributions. The density function of each data point 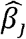 is given by

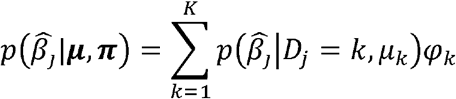

where ***μ* =** {*μ*_0_,…, *μ*_*k*_} and ***φ* =** *φ*_0_,…, *φ*_*k*_}. The log-likelihood function of all beta coefficients 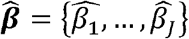 is given by

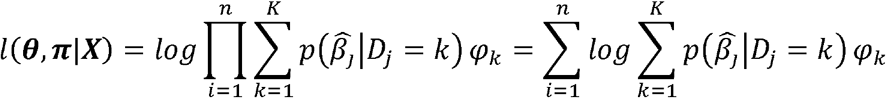

Because there is no closed-form solution of the maximum likelihood estimators (MLE) of the parameters, the expectation–maximization (EM) algorithm is employed to find a numeric solution. The EM algorithm is an iterative method that is described in detail in the next section.

### Parameter estimation by expectation maximization (EM) algorithm

Given a fixed number of signal clusters *K*, we employ the EM algorithm to estimate the 2*K* parameters {(*μ*_*k*_,*φ*_*k*_) ❘ *k* =1, …,*K*}because *φ*_*k*_ are sum to 1, that is,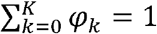. The algorithm undergoes three steps: (1) an initialization step to set the initial values of the parameters; (2) an expectation step that computes the expected value of the log-likelihood; and (3) a maximization step to update the parameters by maximizing the expectation of the log-likelihood. (4) determine the number of clusters K^*^ by the minimum BIC with 2≤K^*^≤7.

#### Initialization

The EM algorithm may converge to a local optimum, and the chance of converging to a global optimum (MLE) depends essentially on the initial values of the parameters.^9^ To minimize the effect of initialization, we set initial values by a crude estimation of the proportion of non-causal variants *φ*_0_ using a variable threshold approach. Suppose the p-values of multiple-variant models of all the rare variants within a region are S = {*p*_*j*_,*j* = 1… *J*}. We define an absolute value of the standardized beta coefficient (Z-score) as 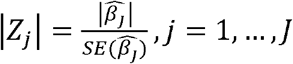. For each p-value, we use a variable threshold approach, that is, *p*_*l*_ is used a threshold, here, *p*_*l*_ ∈ *S*. We meta-analyze the ❘*Z*_*j*_❘ whose corresponding p-value *p*_*j*_ ≤ *p*_*l*_ by the weighted sum of the Z-scores method: 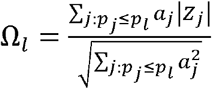, where *a*_*j*_ is a pre-specified weight for j^th^ absolute value of standardized beta.

Different weighting schemes can be used. An appropriate choice of weight may improve the accuracy of the estimation of *φ*_0_. We select two weighting schemes: equal weight (*a*_*j*_ = 1) and weight as the inverse of the estimated standard error 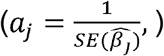. Given a specified weighting scheme, we define an optimal threshold as 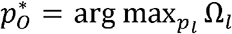. Then, we define a rare variant as a preliminary signal variant if the corresponding p-value 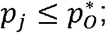; otherwise, the variant is defined as a preliminary non-signal variant. The initial value of *φ*_0_ can be calculated by 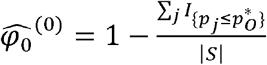. |*S*| is the number of p-values in the set. The preliminary signal variants are then clustered by K-means analysis. The remaining mixing proportion parameters 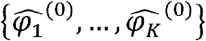 are calculated as 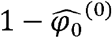 multiplied by the proportion of variants in the K signal clusters from the K-means clustering. The initial values of the means of the signal clusters 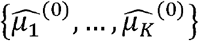 are set as the cluster means from the K-means analysis.

We add a constraint that the preliminary signal variants with opposite effect directions cannot be assigned to the same cluster by the K-means method. That is, the variants with positive beta coefficients and negative ones are split into two separate clusters by the K-means method. Specifically, *K* = *K*+ + *K*-, where *K*+ and *K*-are defined as the number of clusters for the variants with positive and negative beta coefficients, respectively. Then we run K-1 combinations ((*K*+ = 1, *K*- = *K* – 1),…, (*K*+ = 1, *K*- = *K* – 1)) of K-means analysis. The optimal cluster partition is identified by minimizing the within-cluster sum of squares (WCSS) across the K-1 combinations:

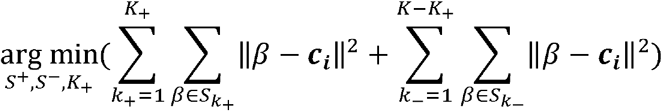

Where 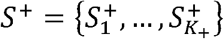 is a partition of the variants with positive effect direction, and 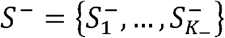 is a partition of variants with a negative effect direction.

#### Expectation step

Because the EM algorithm is an iterative method, we designate 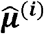 and 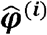 as the values of 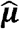 and 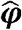 for i^th^ iteration, respectively. For simplicity, we use the 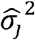 value estimated from the multiple-variant models, and therefore, we do not update 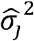 in the EM algorithm. Then we define the posterior probabilities of *Dj* by 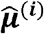 and 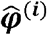 for each iteration

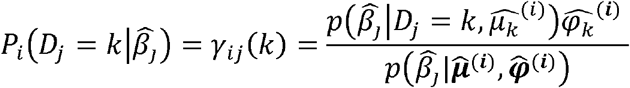

The expectation of the log-likelihood w.r.t current posterior probabilities of *Dj* given 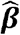 is

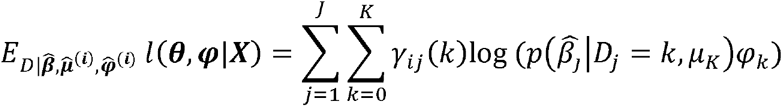

Note that 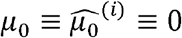.

#### Maximization step

To maximize parameters, we take partial derivatives of 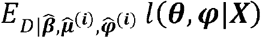 w.r.t each parameter of ***μ*** = {*μ*_1_,…, *μ*_K_}and ***φ*** ={*φ*_1_,…, *φ*_*K*_} and set to 0. That is

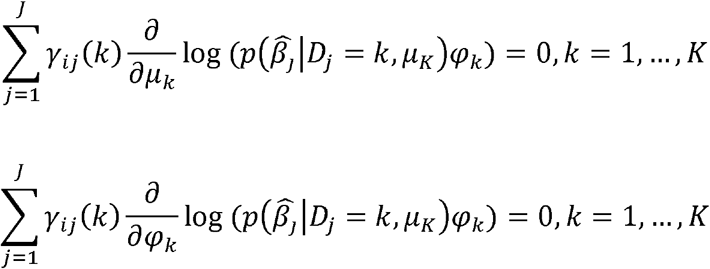

By solving the above equations and adding the constraint of 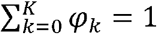, we have

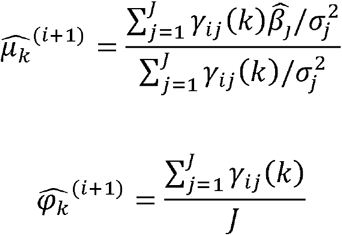

The expectation and maximization steps are implemented iteratively. The algorithm stops when |*l* (θ, π |*X*)^(*i*+1)^ - *l* (θ, π |*X*) ^(*i*)^ |≤ *ε* for with a small positive number *ε*. We set *ε* = 0.001.

For a given K^*^, we choose the clustering results with a higher likelihood from the two sets of initialization values. We adopt the Bayesian information criterion (BIC) to determine the optimal number of clusters with 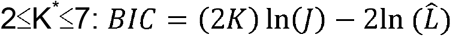, where *J* is the number of variants and 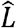 is the maximized value of the likelihood function.

### A simulation study

We simulated a continuous phenotype by the following model:

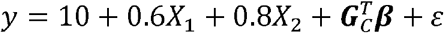

where 10 is the intercept, *X*_1_ ∼ *N*(0,1), *X*_2_ ∼ *Binomial* (0.5) and *ε* ∼ *N* (0,0.49). 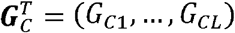 is a vector that includes the genetic coding for *L* randomly chosen causal rare variants in the simulated region. **β** = (β_1_,…, β_*L*_)^*T*^is a vector of true beta effects for the selected causal variants. The beta effect of rare variant *i* is based on *R*^2^and **ω**. *R*^2^is the proportion of variance explained by all of the causal rare variants for a continuous trait. **ω** = {ω_1_,…, ω_*L*_}is the proportion of the beta effects of each causal rare variant. The elements of **ω**are randomly assigned to each causal rare variant.

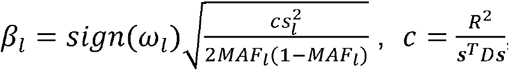, where *D* is the correlation matrix between causal variants, and ***s*** = (*s*_1_,…,*s*_*l*_), where 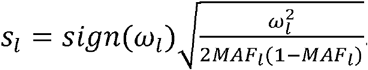. The proportion of variance (R^2^) explained by the causal rare variants was set to be 3% for the continuous phenotype. We also applied a cutoff of 80% quantile to the simulated continuous phenotype with R^2^ = 3% to obtain a binary phenotype. The variance explained by *X*_1_,*X*_2_ and random error was around 36%, 16%, and 45% respectively.

We conduct a simulation study of 1,000 replicates to evaluate the performance of the proposed clustering method. We generate 60 rare variants with a minor allele frequency (MAF) in 0.002≤MAF<0.01. It has been shown that rare variants display mild linkage disequilibrium (LD) between each other.^10^ LD may affect the performance of the clustering because the GMM model assumes the data points are independent. To evaluate the effect of LD on rare variant clustering, we perform simulations to generate rare variants under two conditions: independent variants and those that display LD using human genome sequence data from the 1000 Genome Project Build 37 as the reference.^11^ For the first condition, we generate genotypes using the PhenotypeSimulator R package.^12^ This package simulates genotypes based on the binomial distribution that does not incorporate LD. Second, we generate genotypes using the sim1000g R package that uses the first 60 rare variants of the B-Cell Translocation Gene 3 (BTG3) gene (19,249 base pair in length, GRCh37/hg19) on chromosome 21 in the human 1000 Genomes Project (Build 37) as the reference. Of note, the choice of the BTG3 gene is arbitrary. The sim1000G package simulates variants for small or large genomic regions or a full chromosome in unrelated individuals or family data. Haplotypes are extracted to compute LD in the simulated genomic regions and to generate new genotype data among individuals. For both situations, i.e., the presence and absence of LD by the two R packages, we simulate 1000 replicates of 5,000, 15,000, 25,000 samples. To compare the effect of sample size on rare variant clustering, we randomly select 5000, 15,000 and 25,000 samples from the underlying population of 250,000 individuals. To understand the LD structure between rare variants generated by the sim1000g package, we calculate an average correlation matrix of the simulated 60 rare variants over the 1000 replicates.

For both the continuous and binary traits, we compare the performance of the clustering methods under six simulation scenarios (**Table 1**). For scenario 1: 1/3 of the variants within the target region is non-causal, 1/3 of the variants are causal with moderate positive effect, and 1/3 of the variants are causal with strong positive effect. The ratio of moderate and strong effects is 1:2. In scenario 2: 1/3 of the variants within the target region are non-causal, 1/3 of the variants are causal with positive effect, and 1/3 of the variants are causal with negative effect. The effect size of each causal variant is the same for this scenario. In scenario 3: 1/4 of the variants within the target region are non-causal, 1/4 of the variants are causal with moderate positive effect, 1/4 of the variants are causal with strong positive effect, and 1/4 of the variants are causal with moderate negative effect. The ratio of the effects is -1:0:1:2 for this scenario. In scenario 4: 1/5 of the variants within the target region is non-causal, 1/5 of the variants are causal with moderate positive effect, 1/5 of the variants are causal with strong positive effect, 1/5 of the variants are causal with moderate negative effect, and 1/5 of the variants are causal with strong negative effect. The ratio of the effects is -2:-1:0:1:2. In scenario 5: 1/2 of the variants are causal with moderate positive effect, and the other 1/2 of the variants are causal with strong positive effect. The ratio of the effects is 1:2. In scenario 6: 1/2 of the variants are causal with positive effect, and the other 1/2 of the variants are causal with negative effect. The ratio of the effects is -1:1. Note that scenarios 5 and 6 are two extreme cases that do not contain a null cluster.

**Table 1.** Summary of six simulation scenarios with 1000 replicates.

|  | Number of groups<br>of rare variants | The ratio of true<br>beta effects<br>between each<br>group | Proportions of the<br>number of rare<br>variants among each<br>group |
| --- | --- | --- | --- |
| Scenario 1 | 3 | 0:1:2 | $\frac{1}{3} : \frac{1}{3} : \frac{1}{3}$ |
| Scenario 2 | 3 | -1:0:1 | $\frac{1}{3} : \frac{1}{3} : \frac{1}{3}$ |
| Scenario 3 | 4 | -1:0:1:2 | $\frac{1}{4} : \frac{1}{4} : \frac{1}{4} : \frac{1}{4}$ |
| Scenario 4 | 5 | -2:-1:0:1:2 | $\frac{1}{5} : \frac{1}{5} : \frac{1}{5} : \frac{1}{5} : \frac{1}{5}$ |
| Scenario 5 | 2 | 1:2 | $\frac{1}{2} : \frac{1}{2}$ |
| Scenario 6 | 2 | -1:1 | $\frac{1}{2} : \frac{1}{2}$ |

We adopt the Adjusted Rand index^13^ (ARI) method to measure the similarity between the true and predicted allocations of clusters. That is, we calculate the average ARI over the 1000 replicates to evaluate the performance of the method. For each scenario, we also calculate the accuracy of the number of clusters K^*^ determination. The accuracy is calculated as

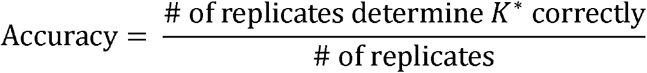

To assess the deviation between mean effects in estimation and the “true” effects from the simulation from clusters, we calculate the mean squared error (MSE) by

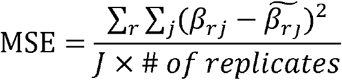

where β_*rj*_ is the true mean of the j^th^ cluster in the r^th^ replicate; and 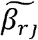 is the corresponding estimated mean based on our clustering algorithm. For both simulation studies and real data analyses, we perform an analysis of variance (ANOVA) to test if the means of betas from different clusters are significantly different. We also apply the Mann-Whitney U test to evaluate if the mean of betas from a null cluster is significantly different from 0.

### Application to exome-wide association and a rare-variant GWAS of blood pressure traits

To demonstrate the proposed clustering method, we apply the method to cluster the rare variants within the significant genes associated with blood pressure (BP) traits. Blood pressure is an inherited trait with an estimated heritability of up to 30-70%.^14^ High blood pressure is an independent risk factor for cardiovascular diseases.^15^ We apply the proposed clustering method to summary statistics obtained from the most updated association studies and meta-analysis of BP traits by Surendran et al.^16^ This study included more than 800,000 individuals from four consortia (CHARGE, CHD Exome+, GoT2D:T2DGenes, ExomeBP) and UK BioBank data.^16^ In this study, an exome-wide association (EWAS) and a rare-variant GWAS (RV-GWAS) using imputed and genotyped single nucleotide variants (SNVs) were conducted to identify common and rare variants, and genes that were associated with three continuous BP traits (systolic blood pressure [SBP], diastolic blood pressure [DBP] and pulse pressure [PP]) and hypertension (HTN) by using both single-variant model and gene-based tests. This large effort validated most of the previously identified BP-associated single variants and genes. In addition, this large study discovered several new SNVs and genes associated with BP traits. ^16^ To cluster rare variants, we consider rare variants in genes that are associated with SBP, DBP, PP, and HTN.^16^ We apply two strategies to cluster rare variants. In the first strategy, we cluster rare variants per gene and trait. In the second strategy, we cluster the combined rare variants per gene.

## RESULTS

### Simulation studies

We first compared clustering methods under different scenarios using independent rare variants generated by the PhenotypeSimulator R package.^12^ We then evaluated the effect of LD on the clustering method for rare variants generated by the sim1000g R package.

#### Simulations without LD structure

The ARI value was largely improved when the sample size was increased, indicating that a large sample size provided a more accurate allocation of the clusters. For example, with a sample size of 5000, the mean ARI value was 0.61 for the combined weighting scheme for a continuous trait under simulation scenario 2. With a sample size of 15,000 and 25,000, the ARI value increased to 0.90 and 0.96, respectively. The ARI values were comparable using three Z-score weighting schemes under each scenario (Supplementary Figures 4 and 5). Simulation scenarios 2 and 6 had higher mean ARI values compared to the other scenarios if other conditions were the same. This observation was as expected because the two signal clusters had opposite effect directions and the differences between the true cluster means of the two signal clusters were larger than those of the other scenarios.

With a pre-specified number of clusters K^*^ ranging from 2 to 7, the accuracy of determining the number of clusters was high (>0.9) for all simulation scenarios when the sample size reached 25000 for a continuous trait. The MSE of the estimates for the true cluster means was reduced with the increase in sample size. The ANOVA test was significant for all replicates under each scenario (P<0.05), indicating that the means of betas from the different clustering were significantly different. The Mann-Whitney test presented non-significant results for all the scenarios (P>0.05), indicating that the rare variants allocated to the null cluster display effect sizes not significantly different from 0.

The ARI value of clustering for binary outcomes was lower than that of continuous outcomes when all the other conditions remained the same (Supplementary Figure 1 and Supplementary Figures 2-11). For example, when there was no LD and the sample size was 15000, the mean ARI value of a continuous trait with the combined weighting was 0.67 under scenario 1. The corresponding mean ARI value of a binary trait was 0.34.

#### Comparison of Simulations with and without LD

We observed a moderate or low LD between the 60 simulated rare variants using the sim1000g package (Supplementary Figure 1). Among a total of 1770 rare variant pairs from the 60 rare variants, 530 pairs (29.9%) displayed a correlation between 0.01 and 0.05, with 73 pairwise displaying correlations > 0.05. The strongest correlation between the two variants was 0.26. The two variants had MAF 0.0031 and 0.0032.

For simulation scenarios 2, 4, and 6, the simulated genotype displaying mostly moderate LD provided comparable mean ARI values, MSE, and accuracy of specification of the number of clusters K^*^ compared to those of independent rare variants (Figure 1, Supplementary Figure 2). For example, the mean ARI values with/without the presence of LD among variants were comparable (0.91 vs. 0.9) in scenario 2 with a sample size of 15,000 for a continuous trait using the combined weighting scheme. The corresponding MSE (0.0023 vs. 0.0022) and accuracy of specification of the number of clusters K^*^ (0.992 vs. 0.995) were similar between the presence and absence of LD structure. For simulation scenarios 1, 3, and 5, the simulated genotype displaying mostly moderate LD provided a lower mean ARI value, larger MSE, and lower accuracy of specification of the number of clusters K^*^ compared to those of independent rare variants (Table 2, Supplementary Tables 1-5, Figure 1, Supplementary Figures 4-5). For example, using the combined weighting scheme, in a sample size of 15,000 with a continuous trait, the average ARI value was 0.53 when the simulated rare variants displayed LD for scenario 1. In contrast, when simulated genotypes were independent, the average ARI value was 0.67, which was about 0.14 higher than the clustering result based on the variants with LD using the same sample size. The corresponding MSE from clustering rare variants without LD structure was 0.0033 (Table 2), while the MSE from cluster rare variants with LD was 0.0043 (Supplementary Table 1). The specification accuracy of the number of K^*^ clusters with the presence of LD (0.715) was lower than that (0.911) with the absence of LD between variants. Comparing the clustering performance of the scenarios between the presence and absence of LD, we observed that the presence of LD had a larger effect on the performance of clustering for scenarios 1, 3, and 5 compared to scenarios 2, 4, and 6. It was likely due to the smaller differences between the true cluster means in scenarios 1, 3, and 5 compared to those in scenarios 2, 4, and 6.

**Figure 1.**
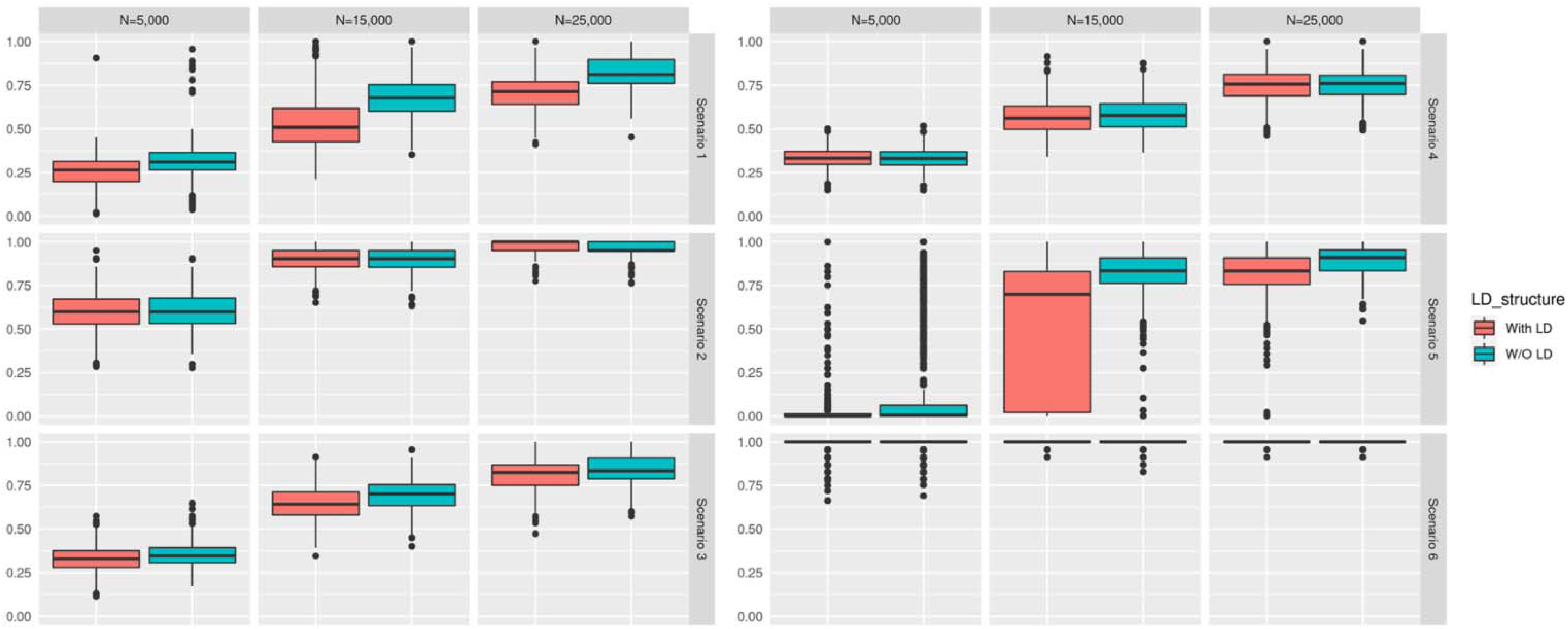
Boxplots of ARI values of simulation scenarios 1, 2, 3, 4, 5 and 6 with combined weighting scheme with/without LD for a continuous trait.

**Table 2.**
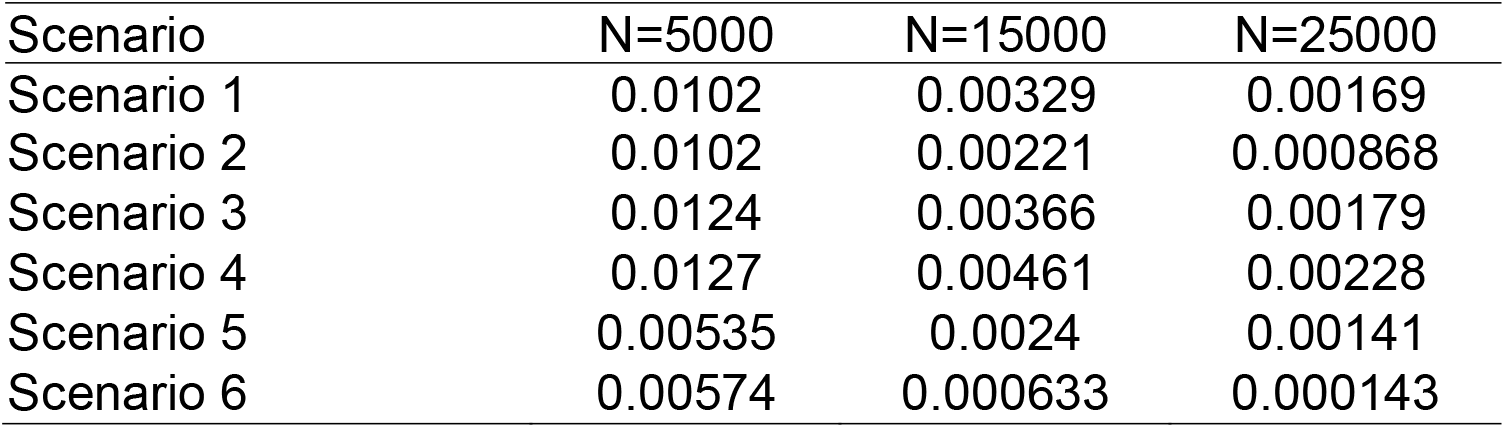
MSE of estimated clusters’ means for a continuous trait with the absence of LD in simulation studies.

### Application to GWAS of BP traits with rare variants

#### Identification of blood pressure trait-associated genes

Using the SKAT test, multiple rare variants (MAF<0.01) were identified for one or more BP traits (P<2.5×10E-6) with four genes (*NPR1, DBH, COL21A1*, and *NOX4*).^8,17^ Low frequency and rare variants in two additional genes of *PLCB3* and *CEP120* were associated with BP traits at MAF<0.05. The six genes harbor different numbers of rare variants. More specifically, *NPR1* included 13 rare variants, *DBH* included 29 rare variants, *COL21A1* included 26 rare variants and *NOX4* included 9 rare variants (Supplementary Table 6). SBP was associated with *NPR1, DBH*, and *PLCB3*; DBP was associated with *DBH* and *PLCB3*; PP was associated with *COL21A1, NOX4*, and *CEP120* due to multiple rare variants in the gene (*P*<2.5E-6). Because gene-based test results were not available for the associations between HTN and these six genes in the GWAS^16^, we defined a signal gene of HTN if any of the six genes contained rare variant(s) (MAF<0.01) with *P*<1E-4 in the single variant-HTN association testing. The three genes of *DBH, NPR1*, and *PLCB3* included rare variant(s) displaying association with HTN at *P*<1E-4. We applied the proposed clustering method to cluster BP-associated rare variants in these genes.

#### Clustering of rare variants

We performed rare variant clustering in eleven gene-trait associations (six genes with four traits): three genes associated with SBP, two genes associated with DBP, three genes associated with PP, and three genes that contain significant rare variants with HTN. On average, a signal gene contains about 20 rare variants. Two to three clusters were identified in each of the eleven gene-trait associations. About 70% of the rare variants were clustered into a null cluster (Table 3 and 4, Supplementary Tables 7-24).

**Table 3.**
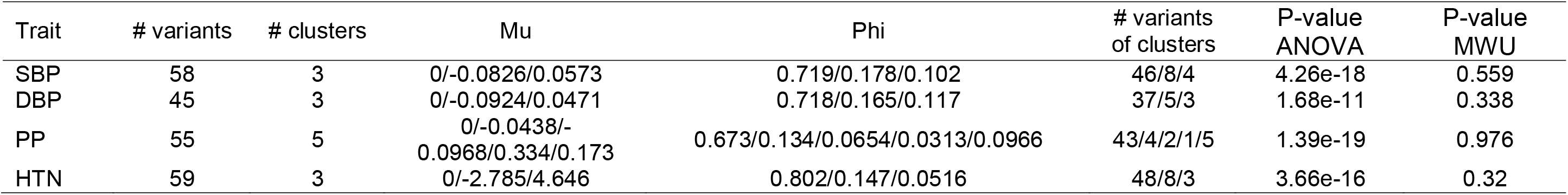
Summary of clustering results for rare variants within the combined signal regions of BP traits.

| Trait | # variants | # clusters | Mu | Phi | # variants<br>of clusters | P-value<br>ANOVA | P-value<br>MWU |
| --- | --- | --- | --- | --- | --- | --- | --- |
| SBP | 58 | 3 | 0/-0.0826/0.0573 | 0.719/0.178/0.102 | 46/8/4 | 4.26e-18 | 0.559 |
| DBP | 45 | 3 | 0/-0.0924/0.0471 | 0.718/0.165/0.117 | 37/5/3 | 1.68e-11 | 0.338 |
| PP | 55 | 5 | 0/-0.0438/-<br>0.0968/0.334/0.173 | 0.673/0.134/0.0654/0.0313/0.0966 | 43/4/2/1/5 | 1.39e-19 | 0.976 |
| HTN | 59 | 3 | 0/-2.785/4.646 | 0.802/0.147/0.0516 | 48/8/3 | 3.66e-16 | 0.32 |

**Table 4.** Summary of clustering results for rare variants within the signal genes of SBP.

| Gene | Chr | Start (bp) | End (bp) | # variants | # clusters | mu | phi | # variants<br>of clusters | P-value<br>_ANOVA | P-value<br>MWU |
| --- | --- | --- | --- | --- | --- | --- | --- | --- | --- | --- |
| DBH | 9 | 136501569 | 136523555 | 27 | 2 | 0/-0.082 | 0.75/0.25 | 21/6 | 5.18e-08 | 0.785 |
| NPR1 | 1 | 153652129 | 153665650 | 13 | 3 | 0/-<br>0.0846/0.164 | 0.727/0.198/0.0753 | 10/2/1 | 6.02e-05 | 0.322 |
| PLCB3 | 11 | 64021930 | 64034975 | 18 | 2 | 0/0.0529 | 0.716/0.284 | 15/3 | 2.33e-05 | 0.934 |

For example, the NPR1 gene contained 12 rare variants. We identified three clusters of rare variants in this gene for the HTN association. The means of standardized beta coefficients (z-scores) were significantly different across the three clusters (ANOVA *P*=0.00028). Of the 12 rare variants, 8 variants were in the null cluster. The rare variants allocated to the null cluster displayed effect sizes (i.e., the standardized beta coefficients) not significantly different from zero (Mann-Whitney U test *P*=0.46). Three variants, including rs140425746, rs61757359, and rs61758562, were grouped into a cluster with an average effect size of -2.05. The rare variant, rs116245325, which displayed the smallest p-value of 1.46E-5 with HTN in single variant analysis, forms a single cluster with an effect size of 4.33. Of note, the clustering pattern in this example was similar to the layout in scenario 2 of the simulation study. That is, the two signal clusters had opposite effect directions. (Supplementary Table 23, 24)

We also conducted clustering analyses on the combined summary data from significant genes that were associated with each of the BP traits. Three genes were associated with PP traits. (Supplementary Table 6) A total of 55 variants were located in these three genes. Of the 55 rare variants within these three genes, we identified five clusters with distinct effect sizes (ANOVA *P*=1.39E-19). (Table 3, Supplementary Tables 12) The null cluster contained 43 out of 55 variants. The effect sizes of rare variants allocated to the null cluster displayed were not significantly different from zero (Mann-Whitney U test *P*=0.976). Two rare variants, rs139341533 and rs56061986, were clustered together with an average effect size of -0.097. Four rare variants, rs2303720, rs114280473, rs189429890, and rs144215891, were assigned to a distinct cluster with an average effect size of -0.0438. A single variant, rs200999181, was recognized as the only variant in the cluster with the strongest effect size of 0.334. Five rare variants, rs201955087, rs115079907, rs76146749, rs200401514, and rs2764043, were grouped into a signal cluster with a moderate positive effect size of 0.173. The observed five-cluster pattern was similar to that in scenario 4 of the simulation study. Of note, scenario 4 included four signal clusters: a cluster with a strong positive effect, a cluster with a moderate positive effect, a cluster with a strong negative effect, and a cluster with a moderate negative effect. For both positive and negative effects, the strong effect sizes were around twice the moderate effect sizes. The variants within the clusters of moderate effect sizes were more than the ones within the clusters of strong effects. The effect size of positive associations was larger than the effect size of negative ones. The running time of the BP variants clustering was around 33 seconds with a core of 4GB of memory.

## DISCUSSION

We proposed a new method to cluster rare variants within signal gene regions associated with disease traits based on summary statistics of variant-trait associations. We performed a comprehensive simulation study to evaluate the performance of the proposed method under different scenarios concerning variant effect direction, LD structure, the number of clusters, and the study sample size. In simulation scenarios 1, 3, and 5, we observed that the proposed method provided a higher ARI value, a lower MSE, and a higher accuracy in the specification of the number of clusters with rare variants in the absence of LD compared to those with low LD given the other conditions are the same. This is expected because the GMM model assumes that the data points (i.e., rare variants) are independent. The ARI value is improved and the MSE value is decreased with an increase in sample size. Among the simulated scenarios, scenario 2 yielded the highest ARI value, smallest MSE, and highest accuracy in the specification K^*^ clusters because this scenario had the largest differences between the true means between clusters.

We also applied the proposed method to cluster rare variants of signal genes associated with BP traits using summary statistics from a large GWAS and meta-analysis of BP traits using around 1.3 million individuals.^8^ We first classified the rare variants in individual genes per trait. To further demonstrate the classification utility, we applied the methods with combined variants in several genes that are associated with the same traits. We found that most of the rare variants within the BP traits-associated genes are grouped into the null cluster, indicating that natural selection is likely the main force in shaping the rare variants in the human genome.^18^ In addition, we found that more rare variants were allocated to clusters with negative means than those with positive means.

A common limitation of gene-based tests is that these methods are not able to cluster possibly neutral, risk, and/or protective rare variants in trait-associated regions. The proposed method may overcome this obstacle by clustering rare variants based on the summary statistics at the single variant level obtained from the published large GWAS and meta-analyses (e.g., > 500,000 samples). By performing the proposed clustering analyses, we can distinguish variants with opposite effects or those with different levels of effect sizes in the target region. In addition, the proposed method is computationally efficient for analyzing large-scale sequencing data.

Previous studies have reported inconsistent views about LD structure among rare variants. Some studies assume the independence between rare variants while others assume a mild correlation between rare variants in evaluating rare variants in association studies with common diseases.^10^ Ignoring LD in rare variants may introduce bias or loss of power in association testing. In this study, we conducted simulation studies using independent rare variants and those simulated based on a genomic region on chromosome 21 in the human 1000 Genomes Project. We found that a large proportion (65%) of these simulated rare variants had no LD (r-squared < 0.01), while 32% displayed low pairwise LD (R^2^ between 0.01 and 0.05), and about 4.5% showed R^2^ in 0.05-0.33. By comparing the performance of the proposed method in clustering independent rare variants and rare variants with LD in simulation studies, we found that the proposed method performs better in clustering independent rare variants when the differences between the true means of clusters were relatively small, even if the multiple-variant model provided beta coefficients accounting for LD between rare variants. In addition, the multiple-variant model requires individual-level genotype data. Extending our algorithm to account for LD using existing summary data from large GWAS will be more cost-effective.

In summary, the proposed clustering algorithm identifies risk and/or protective rare variants of distinct magnitudes according to summary statistics of SNP-trait associations. The proposed method can be easily applied to summary statistics from emerging large-scale rare variants GWAS to identify and group trait-associated rare variants into null and signal groups of discrete effect magnitudes. Therefore, this proposed method may facilitate the identification of potentially causal rare variant clusters in genomic regions and ultimately help understand the genetic architecture underlying human complex traits for the discovery of drug targets and the design of gene therapy.

## Data Availability

All data produced in the present study are available upon reasonable request to the authors

