## Supplemental tables and figures for "CLUSTERING OF RARE VARIANTS FOR CAUSAL VARIANTS IDENTIFICATION AND EFFECT DIRECTION CLASSIFICATION"

Sun et al.

**SUPPLEMENTARY MATERIALS**

**Supplementary Tables**

**Supplementary Table 1** MSE of estimated clusters' means for a continuous trait with the presence of LD in simulation studies

| Scenario | N=5000 | N=15000 | N=25000 |
| --- | --- | --- | --- |
| Scenario 1 | 0.0102 | 0.00429 | 0.00225 |
| Scenario 2 | 0.0118 | 0.00226 | 7.00E-04 |
| Scenario 3 | 0.014 | 0.00446 | 0.0022 |
| Scenario 4 | 0.0148 | 0.00557 | 0.00268 |
| Scenario 5 | 0.00405 | 0.00282 | 0.00177 |
| Scenario 6 | 0.0061 | 0.00042 | 0.000116 |

**Supplementary Table 2** Accuracy of number of clusters specification for a continuous trait with the absence of LD in simulation studies

| Scenario | N=5000 | N=15000 | N=25000 |
| --- | --- | --- | --- |
| Scenario 1 | 0.132 | 0.911 | 0.98 |
| Scenario 2 | 0.99 | 0.995 | 0.991 |
| Scenario 3 | 0.093 | 0.994 | 0.995 |
| Scenario 4 | 0.001 | 0.691 | 0.993 |
| Scenario 5 | 0.737 | 0.895 | 0.935 |
| Scenario 6 | 0.788 | 0.91 | 0.954 |

**Supplementary Table 3** Accuracy of number of clusters specification for a continuous trait with the presence of LD in simulation studies

| Scenario | N=5000 | N=15000 | N=25000 |
| --- | --- | --- | --- |
| Scenario 1 | 0.034 | 0.715 | 0.946 |
| Scenario 2 | 0.995 | 0.992 | 0.993 |
| Scenario 3 | 0.031 | 0.965 | 0.995 |
| Scenario 4 | 0 | 0.596 | 0.977 |
| Scenario 5 | 0.585 | 0.828 | 0.874 |
| Scenario 6 | 0.789 | 0.906 | 0.934 |

**Supplementary Table 4** Accuracy of number of clusters specification for a binary trait with the absence of LD in simulation studies

| Scenario | N=5000 | N=15000 | N=25000 |
| --- | --- | --- | --- |
| Scenario 1 | 0.023 | 0.331 | 0.755 |
| Scenario 2 | 0.581 | 0.989 | 0.984 |
| Scenario 3 | 0.003 | 0.239 | 0.784 |
| Scenario 4 | 0 | 0 | 0.011 |
| Scenario 5 | 0.726 | 0.82 | 0.809 |
| Scenario 6 | 0.722 | 0.775 | 0.861 |

**Supplementary Table 5** Accuracy of number of clusters specification for a binary trait with the presence of LD in simulation studies

| Scenario | N=5000 | N=15000 | N=25000 |
| --- | --- | --- | --- |
| Scenario 1 | 0.012 | 0.126 | 0.445 |
| Scenario 2 | 0.507 | 0.983 | 0.985 |
| Scenario 3 | 0 | 0.153 | 0.668 |
| Scenario 4 | 0 | 0 | 0.009 |
| Scenario 5 | 0.696 | 0.792 | 0.839 |
| Scenario 6 | 0.728 | 0.781 | 0.821 |

**Supplementary Table 6** Summary of associations between genes and blood pressure traits in real data analysis

|  | Chr | Start pos | End pos | Number of rare variants | P-value | | |
| --- | --- | --- | --- | --- | --- | --- | --- |
|  |  |  |  |  | SBP | DBP | PP |
| CEP120 | 5 | 122682248 | 122758665 | 21 | - | - | 8.25E-31 |
| COL21A1 | 6 | 55923968 | 56047400 | 26 | - | - | 2.3E-29 |
| DBH | 9 | 136501569 | 136523555 | 29 | 8.85E-22 | 1.77E-32 | - |
| NOX4 | 11 | 89069094 | 89224387 | 9 | - | - | 1.85E-23 |
| NPR1 | 1 | 153652129 | 153665650 | 13 | 2.72E-10 | - | - |
| PLCB3 | 11 | 64021930 | 64034975 | 15 | 4.97E-13 | 4.27E-10 | - |

**Supplementary Table 7** Summary of clustering results for rare variants within signal genes of DBP

| Gene | Chr | Start (bp) | End (bp) | # variants | # clusters | mu | phi | # variants of clusters | P-value _ANOVA | P-value  MWU |
| --- | --- | --- | --- | --- | --- | --- | --- | --- | --- | --- |
| DBH | 9 | 136501569 | 136523555 | 27 | 2 | 0/-0.0927 | 0.786/0.214 | 22/5 | 9.97e-07 | 0.0391 |
| PLCB3 | 11 | 64021930 | 64034975 | 18 | 2 | 0/0.0473 | 0.69/0.31 | 14/4 | 0.000166 | 0.626 |

**Supplementary Table 8** Summary of clustering results for rare variants within signal genes of PP

| Gene | Chr | Start (bp) | End (bp) | # variants | # clusters | mu | phi | # variants of clusters | P-value _ANOVA | P-value  MWU |
| --- | --- | --- | --- | --- | --- | --- | --- | --- | --- | --- |
| CEP120 | 5 | 122682248 | 122758665 | 21 | 2 | 0/-0.0435 | 0.673/0.327 | 18/3 | 9.98e-08 | 0.196 |
| COL21A1 | 6 | 55923968 | 56047400 | 24 | 3 | 0/0.335/0.171 | 0.761/0.0493/0.19 | 19/1/4 | 1.14e-09 | 0.312 |
| NOX4 | 11 | 89069094 | 89224387 | 10 | 2 | 0/-0.0962 | 0.605/0.395 | 7/3 | 0.00125 | 0.937 |

**Supplementary Table 9** Summary of clustering results for rare variants within signal genes of HTN

| Gene | Chr | Start (bp) | End (bp) | #  variants | #  clusters | mu | phi | # variants of clusters | P-value _ANOVA | P-value  MWU |
| --- | --- | --- | --- | --- | --- | --- | --- | --- | --- | --- |
| DBH | 9 | 136501569 | 136523555 | 29 | 2 | 0/-2.612 | 0.712/0.288 | 21/8 | 2.15e-07 | 0.495 |
| NPR1 | 1 | 153652129 | 153665650 | 12 | 3 | 0/-2.053/4.334 | 0.667/0.253/0.079 | 8/3/1 | 0.000282 | 0.461 |
| PLCB3 | 11 | 64021930 | 64034975 | 18 | 2 | 0/4.808 | 0.887/0.113 | 16/2 | 1.27e-05 | 0.433 |

**Supplementary Table 10** Clustering results of rare variants within the combined signal region of SBP

| rsID | Chr | Position | Major allele | Minor allele | MAF | Effect | SE | P-value | Cluster membership | Cluster mean |
| --- | --- | --- | --- | --- | --- | --- | --- | --- | --- | --- |
| rs76856960 | 9 | 136501569 | g | a | 0.0046 | 0.00322 | 0.0138 | 0.8151 | 1 | 0 |
| rs143544421 | 9 | 136501599 | g | a | 5e-04 | 0.123 | 0.156 | 0.4303 | 1 | 0 |
| rs78445536 | 9 | 136501746 | g | a | 5e-04 | -0.0666 | 0.0572 | 0.2443 | 1 | 0 |
| rs146922432 | 9 | 136501768 | g | a | 3e-04 | -0.0174 | 0.144 | 0.9042 | 1 | 0 |
| rs76819676 | 9 | 136507375 | g | a | 2e-04 | -0.0232 | 0.111 | 0.8347 | 1 | 0 |
| rs200430427 | 9 | 136507456 | c | t | 4e-04 | -0.0581 | 0.154 | 0.706 | 1 | 0 |
| rs143535251 | 9 | 136507474 | c | t | 0.0013 | 0.0197 | 0.0248 | 0.4267 | 1 | 0 |
| rs200628504 | 9 | 136507528 | c | t | 4e-04 | 0.0516 | 0.128 | 0.6857 | 1 | 0 |
| rs5321 | 9 | 136507559 | g | c | 5e-04 | 0.0625 | 0.069 | 0.3652 | 1 | 0 |
| rs199734841 | 9 | 136507586 | g | c | 4e-04 | -0.176 | 0.2 | 0.3791 | 1 | 0 |
| rs13306301 | 9 | 136508640 | g | a | 1e-04 | -0.549 | 0.282 | 0.05178 | 1 | 0 |
| rs5324 | 9 | 136508658 | g | a | 6e-04 | -0.0219 | 0.0384 | 0.5682 | 1 | 0 |
| rs145655199 | 9 | 136508682 | g | a | 0.001 | -0.0173 | 0.18 | 0.9233 | 1 | 0 |
| rs201681337 | 9 | 136508691 | g | a | 0.002 | 0.0305 | 0.0754 | 0.6864 | 1 | 0 |
| rs75215331 | 9 | 136513028 | c | t | 0.0034 | 0.0146 | 0.0168 | 0.3834 | 1 | 0 |
| rs41316996 | 9 | 136521654 | g | a | 0.0032 | 0.019 | 0.0145 | 0.1918 | 1 | 0 |
| rs144040856 | 9 | 136521738 | g | a | 3e-04 | -0.0756 | 0.135 | 0.5758 | 1 | 0 |
| rs141021210 | 9 | 136521751 | a | g | 0 | -0.131 | 0.997 | 0.8956 | 1 | 0 |
| rs201973877 | 9 | 136522317 | t | c | 0.0003 | -0.0251 | 0.0565 | 0.6566 | 1 | 0 |
| rs75512464 | 9 | 136523487 | a | t | 0.002 | -0.0264 | 0.0236 | 0.2621 | 1 | 0 |
| rs76316834 | 9 | 136523555 | g | a | 4e-04 | 0.0484 | 0.13 | 0.709 | 1 | 0 |
| rs56019647 | 1 | 153652129 | c | t | 1e-04 | -0.113 | 0.165 | 0.4947 | 1 | 0 |
| rs28730726 | 1 | 153653757 | g | c | 0.0016 | -0.0175 | 0.0432 | 0.6842 | 1 | 0 |
| rs199612927 | 1 | 153654234 | g | a | 2e-04 | 0.0688 | 0.0871 | 0.4298 | 1 | 0 |
| rs140425746 | 1 | 153655966 | g | a | 2e-04 | -0.0333 | 0.154 | 0.8287 | 1 | 0 |
| rs201746049 | 1 | 153656216 | a | g | 0.0003 | -0.033 | 0.195 | 0.8658 | 1 | 0 |
| rs115938602 | 1 | 153656228 | c | a | 0 | -0.139 | 0.29 | 0.6323 | 1 | 0 |
| rs149202797 | 1 | 153658306 | c | a | 2e-04 | -0.0203 | 0.136 | 0.8808 | 1 | 0 |
| rs116775696 | 1 | 153659550 | a | g | 0 | -0.333 | 0.458 | 0.4672 | 1 | 0 |
| rs139174442 | 1 | 153660154 | c | g | 10.0e-05 | 0.297 | 0.35 | 0.3973 | 1 | 0 |
| rs201787421 | 1 | 153660625 | g | a | 2e-04 | -0.68 | 0.316 | 0.0312 | 1 | 0 |
| rs200996360 | 11 | 640253 | c | g | 0.0003 | -0.03 | 0.0527 | 0.5689 | 1 | 0 |
| rs111961110 | 11 | 64021930 | a | g | 0.0003 | -0.14 | 0.207 | 0.4974 | 1 | 0 |
| rs144191345 | 11 | 64022785 | c | t | 2e-04 | -0.0212 | 0.108 | 0.8442 | 1 | 0 |
| rs199645363 | 11 | 64022873 | c | t | 3e-04 | 0.0912 | 0.101 | 0.3668 | 1 | 0 |
| rs138400940 | 11 | 64022905 | c | t | 1e-04 | -0.381 | 0.576 | 0.5084 | 1 | 0 |
| rs188572550 | 11 | 64026357 | g | a | 0 | 0.801 | 0.704 | 0.2551 | 1 | 0 |
| rs201842672 | 11 | 64027567 | g | t | 0.0038 | -0.0627 | 0.0711 | 0.3777 | 1 | 0 |
| rs200923408 | 11 | 64028916 | g | a | 3e-04 | -0.538 | 0.235 | 0.02196 | 1 | 0 |
| rs200263631 | 11 | 64031571 | t | c | 0.0003 | 0.0847 | 0.0703 | 0.2283 | 1 | 0 |
| rs79573066 | 11 | 64032784 | g | a | 0.0036 | 0.0262 | 0.0696 | 0.7066 | 1 | 0 |
| rs61757725 | 11 | 64033360 | g | t | 0.009 | 0.0786 | 0.0599 | 0.1896 | 1 | 0 |
| rs148059922 | 11 | 64033391 | g | c | 4e-04 | -0.0149 | 0.131 | 0.9093 | 1 | 0 |
| rs201342752 | 11 | 64033986 | c | t | 0.002 | -0.0891 | 0.0706 | 0.2068 | 1 | 0 |
| rs141163685 | 11 | 64034734 | g | t | 0.0031 | 0.249 | 0.113 | 0.02777 | 1 | 0 |
| rs113554478 | 11 | 64034975 | g | a | 0.0011 | 0.00854 | 0.102 | 0.9333 | 1 | 0 |
| rs3025380 | 9 | 136501756 | g | c | 0.0045 | -0.0884 | 0.0119 | 1.096e-13 | 2 | -0.0826 |
| rs74853476 | 9 | 136501834 | t | c | 0.0021 | -0.0774 | 0.0181 | 1.973e-05 | 2 | -0.0826 |
| rs142383279 | 9 | 136507332 | g | a | 0.0018 | -0.0652 | 0.0194 | 0.0007811 | 2 | -0.0826 |
| rs145059403 | 9 | 136507425 | g | a | 0.001 | -0.102 | 0.0261 | 9.144e-05 | 2 | -0.0826 |
| rs148439785 | 9 | 136521726 | g | a | 8e-04 | -0.0595 | 0.0313 | 0.05706 | 2 | -0.0826 |
| rs151228388 | 9 | 136522272 | a | g | 0.0013 | -0.317 | 0.102 | 0.001909 | 2 | -0.0826 |
| rs61757359 | 1 | 153658297 | g | a | 0.0034 | -0.0819 | 0.014 | 4.487e-09 | 2 | -0.0826 |
| rs61758562 | 1 | 153659131 | g | a | 5e-04 | -0.133 | 0.0495 | 0.007185 | 2 | -0.0826 |
| rs116245325 | 1 | 153665650 | c | t | 8e-04 | 0.166 | 0.0287 | 7.686e-09 | 3 | 0.0573 |
| rs117874826 | 11 | 64027666 | a | c | 0.0138 | 0.0467 | 0.00729 | 1.535e-10 | 3 | 0.0573 |
| rs145502455 | 11 | 64031030 | g | a | 0.0054 | 0.0723 | 0.0118 | 8.563e-10 | 3 | 0.0573 |
| rs142330950 | 11 | 64032945 | g | t | 0.0021 | 0.0456 | 0.0182 | 0.01236 | 3 | 0.0573 |

**Supplementary Table 11** Clustering results of rare variants within the combined signal region of DBP

| rsID | Chr | Position | Major allele | Minor allele | MAF | Effect | SE | P-value | Cluster membership | Cluster mean |
| --- | --- | --- | --- | --- | --- | --- | --- | --- | --- | --- |
| rs76856960 | 9 | 136501569 | g | a | 0.0046 | -0.00876 | 0.0138 | 0.5248 | 1 | 0 |
| rs143544421 | 9 | 136501599 | g | a | 5e-04 | 0.0145 | 0.156 | 0.9262 | 1 | 0 |
| rs78445536 | 9 | 136501746 | g | a | 5e-04 | -0.0651 | 0.0572 | 0.255 | 1 | 0 |
| rs146922432 | 9 | 136501768 | g | a | 3e-04 | -0.125 | 0.144 | 0.3888 | 1 | 0 |
| rs76819676 | 9 | 136507375 | g | a | 2e-04 | 0.0357 | 0.111 | 0.7477 | 1 | 0 |
| rs200430427 | 9 | 136507456 | c | t | 4e-04 | 0.0385 | 0.154 | 0.8029 | 1 | 0 |
| rs143535251 | 9 | 136507474 | c | t | 0.0013 | 0.00881 | 0.0248 | 0.7226 | 1 | 0 |
| rs200628504 | 9 | 136507528 | c | t | 4e-04 | -0.081 | 0.128 | 0.5253 | 1 | 0 |
| rs5321 | 9 | 136507559 | g | c | 5e-04 | 0.0583 | 0.069 | 0.3984 | 1 | 0 |
| rs199734841 | 9 | 136507586 | g | c | 4e-04 | -0.225 | 0.2 | 0.2601 | 1 | 0 |
| rs13306301 | 9 | 136508640 | g | a | 1e-04 | -0.753 | 0.283 | 0.007722 | 1 | 0 |
| rs5324 | 9 | 136508658 | g | a | 6e-04 | -0.0431 | 0.0385 | 0.2625 | 1 | 0 |
| rs145655199 | 9 | 136508682 | g | a | 0.001 | 0.129 | 0.18 | 0.473 | 1 | 0 |
| rs201681337 | 9 | 136508691 | g | a | 0.002 | 0.0397 | 0.0755 | 0.5993 | 1 | 0 |
| rs75215331 | 9 | 136513028 | c | t | 0.0034 | 0.00505 | 0.0168 | 0.7633 | 1 | 0 |
| rs41316996 | 9 | 136521654 | g | a | 0.0032 | 0.00855 | 0.0146 | 0.557 | 1 | 0 |
| rs148439785 | 9 | 136521726 | g | a | 8e-04 | -0.0323 | 0.0313 | 0.3019 | 1 | 0 |
| rs144040856 | 9 | 136521738 | g | a | 3e-04 | -0.217 | 0.135 | 0.1089 | 1 | 0 |
| rs141021210 | 9 | 136521751 | a | g | 0 | -1.023 | 0.995 | 0.3037 | 1 | 0 |
| rs201973877 | 9 | 136522317 | t | c | 0.0003 | -0.0286 | 0.0566 | 0.6134 | 1 | 0 |
| rs75512464 | 9 | 136523487 | a | t | 0.002 | -0.025 | 0.0236 | 0.2893 | 1 | 0 |
| rs76316834 | 9 | 136523555 | g | a | 4e-04 | 0.015 | 0.13 | 0.9078 | 1 | 0 |
| rs200996360 | 11 | 640253 | c | g | 0.0003 | 0.0237 | 0.0528 | 0.653 | 1 | 0 |
| rs111961110 | 11 | 64021930 | a | g | 0.0003 | 0.00639 | 0.207 | 0.9754 | 1 | 0 |
| rs144191345 | 11 | 64022785 | c | t | 2e-04 | 0.0759 | 0.108 | 0.4822 | 1 | 0 |
| rs199645363 | 11 | 64022873 | c | t | 3e-04 | 0.0863 | 0.101 | 0.3935 | 1 | 0 |
| rs138400940 | 11 | 64022905 | c | t | 1e-04 | -0.858 | 0.576 | 0.1362 | 1 | 0 |
| rs188572550 | 11 | 64026357 | g | a | 0 | 0.624 | 0.704 | 0.3759 | 1 | 0 |
| rs201842672 | 11 | 64027567 | g | t | 0.0038 | -0.0781 | 0.0711 | 0.2721 | 1 | 0 |
| rs200923408 | 11 | 64028916 | g | a | 3e-04 | -0.194 | 0.235 | 0.4077 | 1 | 0 |
| rs200263631 | 11 | 64031571 | t | c | 0.0003 | 0.121 | 0.0704 | 0.08613 | 1 | 0 |
| rs79573066 | 11 | 64032784 | g | a | 0.0036 | 0.0198 | 0.0697 | 0.7768 | 1 | 0 |
| rs61757725 | 11 | 64033360 | g | t | 0.009 | 0.0537 | 0.06 | 0.3709 | 1 | 0 |
| rs148059922 | 11 | 64033391 | g | c | 4e-04 | -0.154 | 0.131 | 0.2409 | 1 | 0 |
| rs201342752 | 11 | 64033986 | c | t | 0.002 | -0.0181 | 0.0706 | 0.7979 | 1 | 0 |
| rs141163685 | 11 | 64034734 | g | t | 0.0031 | 0.214 | 0.113 | 0.05838 | 1 | 0 |
| rs113554478 | 11 | 64034975 | g | a | 0.0011 | 0.0951 | 0.102 | 0.3518 | 1 | 0 |
| rs3025380 | 9 | 136501756 | g | c | 0.0045 | -0.103 | 0.0119 | 4.287e-18 | 2 | -0.0924 |
| rs74853476 | 9 | 136501834 | t | c | 0.0021 | -0.0954 | 0.0182 | 1.529e-07 | 2 | -0.0924 |
| rs142383279 | 9 | 136507332 | g | a | 0.0018 | -0.0701 | 0.0194 | 0.0003159 | 2 | -0.0924 |
| rs145059403 | 9 | 136507425 | g | a | 0.001 | -0.0829 | 0.0262 | 0.001551 | 2 | -0.0924 |
| rs151228388 | 9 | 136522272 | a | g | 0.0013 | -0.226 | 0.102 | 0.02711 | 2 | -0.0924 |
| rs117874826 | 11 | 64027666 | a | c | 0.0138 | 0.041 | 0.00731 | 2.069e-08 | 3 | 0.0471 |
| rs145502455 | 11 | 64031030 | g | a | 0.0054 | 0.0668 | 0.0118 | 1.619e-08 | 3 | 0.0471 |
| rs142330950 | 11 | 64032945 | g | t | 0.0021 | 0.0367 | 0.0182 | 0.04445 | 3 | 0.0471 |

**Supplementary Table 12** Clustering results of rare variants within the combined signal region of PP

| rsID | Chr | Position | Major allele | Minor allele | MAF | Effect | SE | P-value | Cluster membership | Cluster mean |
| --- | --- | --- | --- | --- | --- | --- | --- | --- | --- | --- |
| rs145436175 | 5 | 122682248 | t | c | 0.0008 | 0.233 | 0.268 | 0.385 | 1 | 0 |
| rs140306974 | 5 | 122685717 | g | a | 0.0011 | 0.0594 | 0.126 | 0.6367 | 1 | 0 |
| rs200061679 | 5 | 122685731 | t | a | 0.0015 | 0.0692 | 0.0388 | 0.07491 | 1 | 0 |
| rs142792779 | 5 | 122708381 | c | t | 3e-04 | -0.0136 | 0.213 | 0.9492 | 1 | 0 |
| rs139865050 | 5 | 122713092 | c | g | 0 | -0.192 | 0.707 | 0.7861 | 1 | 0 |
| rs74938108 | 5 | 122713159 | c | t | 0.0016 | -0.0257 | 0.0905 | 0.7765 | 1 | 0 |
| rs201600892 | 5 | 122713191 | c | g | 10.0e-05 | 0.0767 | 0.447 | 0.8637 | 1 | 0 |
| rs61744334 | 5 | 122714044 | t | c | 0.0006 | -0.0201 | 0.0684 | 0.7689 | 1 | 0 |
| rs144490830 | 5 | 122714104 | g | c | 2e-04 | 0.37 | 0.179 | 0.03908 | 1 | 0 |
| rs147277049 | 5 | 122720724 | t | c | 0.0006 | -0.0181 | 0.036 | 0.6152 | 1 | 0 |
| rs200450605 | 5 | 122725693 | g | a | 0.0017 | 0.016 | 0.0229 | 0.4841 | 1 | 0 |
| rs201571160 | 5 | 122725754 | c | g | 0.0003 | 0.166 | 0.316 | 0.6004 | 1 | 0 |
| rs114281792 | 5 | 122725761 | t | c | 0.0067 | -0.00233 | 0.0098 | 0.8124 | 1 | 0 |
| rs61747983 | 5 | 122725768 | g | a | 1e-04 | 0.0156 | 0.112 | 0.8895 | 1 | 0 |
| rs147273517 | 5 | 122748194 | t | c | 10.0e-05 | 0.36 | 0.162 | 0.02599 | 1 | 0 |
| rs202103949 | 5 | 122748198 | t | c | 0.0004 | -0.029 | 0.121 | 0.8106 | 1 | 0 |
| rs199793672 | 5 | 122758665 | t | c | 10.0e-05 | -0.479 | 0.503 | 0.3404 | 1 | 0 |
| rs200478915 | 6 | 55923968 | g | c | 8e-04 | -0.0449 | 0.0323 | 0.1652 | 1 | 0 |
| rs200564236 | 6 | 55925008 | g | a | 8e-04 | -0.0212 | 0.162 | 0.8957 | 1 | 0 |
| rs199722485 | 6 | 55925588 | t | g | 0.0005 | -0.0128 | 0.106 | 0.9038 | 1 | 0 |
| rs200674177 | 6 | 55925689 | g | a | 2e-04 | -0.0981 | 0.189 | 0.6046 | 1 | 0 |
| rs201839603 | 6 | 55925762 | a | c | 10.0e-05 | 0.0193 | 0.577 | 0.9733 | 1 | 0 |
| rs9464337 | 6 | 55925801 | g | t | 0.0054 | 0.000204 | 0.0116 | 0.986 | 1 | 0 |
| rs201892311 | 6 | 55926464 | g | c | 1e-04 | 0.426 | 0.707 | 0.547 | 1 | 0 |
| rs199910287 | 6 | 55935556 | g | a | 6e-04 | 0.054 | 0.0445 | 0.2255 | 1 | 0 |
| rs191626317 | 6 | 55939059 | g | t | 0.002 | 0.0424 | 0.102 | 0.6783 | 1 | 0 |
| rs75605879 | 6 | 55966311 | a | g | 10.0e-05 | -0.147 | 0.107 | 0.169 | 1 | 0 |
| rs202115077 | 6 | 55988871 | g | t | 0.002 | 0.00866 | 0.0186 | 0.6409 | 1 | 0 |
| rs201267383 | 6 | 56006611 | t | c | 0.0003 | -0.077 | 0.0817 | 0.3461 | 1 | 0 |
| rs35583895 | 6 | 56006732 | a | g | 0.0081 | 0.00138 | 0.0479 | 0.9771 | 1 | 0 |
| rs200361985 | 6 | 56029264 | g | a | 0.002 | 0.0616 | 0.0546 | 0.2593 | 1 | 0 |
| rs142653960 | 6 | 56035494 | c | t | 4e-04 | -0.0656 | 0.0525 | 0.2115 | 1 | 0 |
| rs199532612 | 6 | 56035853 | a | t | 0.0017 | -0.0186 | 0.0217 | 0.3925 | 1 | 0 |
| rs200708113 | 6 | 56035881 | c | t | 5e-04 | -0.0112 | 0.0509 | 0.8259 | 1 | 0 |
| rs202026963 | 6 | 56035909 | g | a | 1e-04 | 0.0193 | 0.242 | 0.9364 | 1 | 0 |
| rs147394600 | 6 | 56047400 | g | a | 1e-04 | -0.159 | 0.164 | 0.331 | 1 | 0 |
| rs115031759 | 11 | 89073269 | g | a | 1e-04 | 0.192 | 0.161 | 0.2323 | 1 | 0 |
| rs201165492 | 11 | 89106599 | c | t | 1e-04 | -0.274 | 0.576 | 0.6345 | 1 | 0 |
| rs149515506 | 11 | 89135492 | a | g | 10.0e-05 | 0.428 | 0.706 | 0.5445 | 1 | 0 |
| rs147350656 | 11 | 89177310 | c | t | 1e-04 | -0.184 | 0.236 | 0.4375 | 1 | 0 |
| rs142433357 | 11 | 89182609 | t | c | 10.0e-05 | -1.231 | 0.707 | 0.08181 | 1 | 0 |
| rs55977241 | 11 | 89182652 | c | t | 0.002 | 0.000803 | 0.109 | 0.9941 | 1 | 0 |
| rs145686545 | 11 | 89224387 | c | t | 2e-04 | 0.162 | 0.202 | 0.4229 | 1 | 0 |
| rs2303720 | 5 | 122682334 | c | t | 0.0291 | -0.0418 | 0.00482 | 4.532e-18 | 2 | -0.0438 |
| rs114280473 | 5 | 122714092 | g | a | 0.0063 | -0.0632 | 0.011 | 8.076e-09 | 2 | -0.0438 |
| rs189429890 | 5 | 122729025 | c | t | 0.0065 | -0.0373 | 0.00999 | 0.0001886 | 2 | -0.0438 |
| rs144215891 | 11 | 89069094 | t | c | 0.0014 | -0.0716 | 0.0302 | 0.01798 | 2 | -0.0438 |
| rs139341533 | 11 | 89182666 | c | a | 0.0043 | -0.0905 | 0.0126 | 8.085e-13 | 3 | -0.0968 |
| rs56061986 | 11 | 89182686 | t | c | 0.0029 | -0.113 | 0.0161 | 2.385e-12 | 3 | -0.0968 |
| rs200999181 | 6 | 55935568 | c | a | 0.0012 | 0.336 | 0.0244 | 3.471e-43 | 4 | 0.334 |
| rs201955087 | 5 | 122727015 | c | t | 0.0013 | 0.274 | 0.106 | 0.009927 | 5 | 0.173 |
| rs115079907 | 6 | 55924005 | c | t | 0.0015 | 0.207 | 0.0248 | 5.573e-17 | 5 | 0.173 |
| rs76146749 | 6 | 55925783 | t | a | 7e-04 | 0.198 | 0.0782 | 0.01144 | 5 | 0.173 |
| rs200401514 | 6 | 55989091 | c | t | 4e-04 | 0.124 | 0.0552 | 0.02532 | 5 | 0.173 |
| rs2764043 | 6 | 56035643 | a | g | 0.0016 | 0.152 | 0.0203 | 7.82e-14 | 5 | 0.173 |

**Supplementary Table 13** Clustering results of rare variants within the combined signal region of HTN

| rsID | Chr | Position | Major allele | Minor allele | MAF | Z value | P-value | Cluster membership | Cluster mean |
| --- | --- | --- | --- | --- | --- | --- | --- | --- | --- |
| rs76856960 | 9 | 136501569 | g | a | 0.0035 | -0.511 | 0.6096 | 1 | 0 |
| rs143544421 | 9 | 136501599 | g | a | 2e-04 | -0.526 | 0.5989 | 1 | 0 |
| rs78445536 | 9 | 136501746 | g | a | 3e-04 | 0.234 | 0.815 | 1 | 0 |
| rs146922432 | 9 | 136501768 | g | a | 1e-04 | 1.257 | 0.2089 | 1 | 0 |
| rs142383279 | 9 | 136507332 | g | a | 0.002 | -1.826 | 0.06781 | 1 | 0 |
| rs76819676 | 9 | 136507375 | g | a | 1e-04 | -0.787 | 0.4313 | 1 | 0 |
| rs143535251 | 9 | 136507474 | c | t | 0.0014 | 0.397 | 0.6916 | 1 | 0 |
| rs200628504 | 9 | 136507528 | c | t | 2e-04 | -0.0675 | 0.9462 | 1 | 0 |
| rs5321 | 9 | 136507559 | g | c | 2e-04 | 0.226 | 0.8212 | 1 | 0 |
| rs199734841 | 9 | 136507586 | g | c | 2e-04 | 0.897 | 0.3697 | 1 | 0 |
| rs13306301 | 9 | 136508640 | g | a | 0 | -1.378 | 0.1683 | 1 | 0 |
| rs5324 | 9 | 136508658 | g | a | 8e-04 | -1.404 | 0.1604 | 1 | 0 |
| rs145655199 | 9 | 136508682 | g | a | 3e-04 | -1.247 | 0.2125 | 1 | 0 |
| rs201681337 | 9 | 136508691 | g | a | 2e-04 | 1.109 | 0.2676 | 1 | 0 |
| rs75215331 | 9 | 136513028 | c | t | 0.0025 | -0.0818 | 0.9348 | 1 | 0 |
| rs41316996 | 9 | 136521654 | g | a | 0.003 | 2.028 | 0.04261 | 1 | 0 |
| rs144040856 | 9 | 136521738 | g | a | 1e-04 | 0.0598 | 0.9523 | 1 | 0 |
| rs141021210 | 9 | 136521751 | a | g | 0 | -0.665 | 0.5059 | 1 | 0 |
| rs201973877 | 9 | 136522317 | t | c | 0.0002 | -0.923 | 0.356 | 1 | 0 |
| rs75512464 | 9 | 136523487 | a | t | 0.0014 | -0.289 | 0.7723 | 1 | 0 |
| rs148806316 | 9 | 136523534 | c | t | 0.0025 | 0.295 | 0.7679 | 1 | 0 |
| rs76316834 | 9 | 136523555 | g | a | 2e-04 | -0.39 | 0.6962 | 1 | 0 |
| rs56019647 | 1 | 153652129 | c | t | 0 | -0.727 | 0.4675 | 1 | 0 |
| rs199612927 | 1 | 153654234 | g | a | 1e-04 | 0.468 | 0.6397 | 1 | 0 |
| rs140425746 | 1 | 153655966 | g | a | 1e-04 | -1.607 | 0.1081 | 1 | 0 |
| rs201746049 | 1 | 153656216 | a | g | 10.0e-05 | -1.254 | 0.21 | 1 | 0 |
| rs115938602 | 1 | 153656228 | c | a | 0 | -1.174 | 0.2405 | 1 | 0 |
| rs149202797 | 1 | 153658306 | c | a | 1e-04 | 0.23 | 0.8182 | 1 | 0 |
| rs61758562 | 1 | 153659131 | g | a | 0.0011 | -1.693 | 0.09054 | 1 | 0 |
| rs116775696 | 1 | 153659550 | a | g | 0 | 0.641 | 0.5213 | 1 | 0 |
| rs139174442 | 1 | 153660154 | c | g | 0 | 0.0493 | 0.9606 | 1 | 0 |
| rs201787421 | 1 | 153660625 | g | a | 1e-04 | -0.287 | 0.7739 | 1 | 0 |
| rs200996360 | 11 | 640253 | c | g | 0.0003 | -0.905 | 0.3653 | 1 | 0 |
| rs111961110 | 11 | 64021930 | a | g | 10.0e-05 | -0.917 | 0.3591 | 1 | 0 |
| rs144191345 | 11 | 64022785 | c | t | 1e-04 | 0.259 | 0.7958 | 1 | 0 |
| rs199645363 | 11 | 64022873 | c | t | 2e-04 | -0.587 | 0.5575 | 1 | 0 |
| rs138400940 | 11 | 64022905 | c | t | 1e-04 | -1.28 | 0.2004 | 1 | 0 |
| rs188572550 | 11 | 64026357 | g | a | 0 | -0.324 | 0.7457 | 1 | 0 |
| rs201842672 | 11 | 64027567 | g | t | 9e-04 | -0.58 | 0.5616 | 1 | 0 |
| rs200923408 | 11 | 64028916 | g | a | 1e-04 | -0.496 | 0.6198 | 1 | 0 |
| rs200263631 | 11 | 64031571 | t | c | 0.0002 | 1.033 | 0.3018 | 1 | 0 |
| rs79573066 | 11 | 64032784 | g | a | 2e-04 | 0.665 | 0.5063 | 1 | 0 |
| rs142330950 | 11 | 64032945 | g | t | 0.0021 | 2.175 | 0.02966 | 1 | 0 |
| rs61757725 | 11 | 64033360 | g | t | 3e-04 | 0.427 | 0.6694 | 1 | 0 |
| rs148059922 | 11 | 64033391 | g | c | 1e-04 | 1.308 | 0.1908 | 1 | 0 |
| rs201342752 | 11 | 64033986 | c | t | 5e-04 | 1.223 | 0.2215 | 1 | 0 |
| rs141163685 | 11 | 64034734 | g | t | 4e-04 | 0.867 | 0.3859 | 1 | 0 |
| rs113554478 | 11 | 64034975 | g | a | 1e-04 | 0.708 | 0.4787 | 1 | 0 |
| rs77273740 | 9 | 136501728 | c | t | 0.0042 | -2.933 | 0.003353 | 2 | -2.785 |
| rs3025380 | 9 | 136501756 | g | c | 0.0045 | -5.297 | 1.176e-07 | 2 | -2.785 |
| rs74853476 | 9 | 136501834 | t | c | 0.0022 | -2.186 | 0.02885 | 2 | -2.785 |
| rs145059403 | 9 | 136507425 | g | a | 0.0012 | -3.468 | 0.0005248 | 2 | -2.785 |
| rs200430427 | 9 | 136507456 | c | t | 1e-04 | -2.306 | 0.02112 | 2 | -2.785 |
| rs148439785 | 9 | 136521726 | g | a | 0.0019 | -2.196 | 0.02806 | 2 | -2.785 |
| rs151228388 | 9 | 136522272 | a | g | 0.0003 | -2.399 | 0.01646 | 2 | -2.785 |
| rs61757359 | 1 | 153658297 | g | a | 0.0036 | -3.719 | 2e-04 | 2 | -2.785 |
| rs116245325 | 1 | 153665650 | c | t | 8e-04 | 4.335 | 1.457e-05 | 3 | 4.646 |
| rs117874826 | 11 | 64027666 | a | c | 0.0308 | 5.712 | 1.119e-08 | 3 | 4.646 |
| rs145502455 | 11 | 64031030 | g | a | 0.0047 | 4.015 | 5.952e-05 | 3 | 4.646 |

**Supplementary Table 14** Clustering results of rare variants within the DBH gene with SBP

| rsID | Chr | Position | Major allele | Minor allele | MAF | Effect | SE | P-value | Cluster membership | Cluster mean |
| --- | --- | --- | --- | --- | --- | --- | --- | --- | --- | --- |
| rs76856960 | 9 | 136501569 | g | a | 0.0046 | 0.00322 | 0.0138 | 0.8151 | 1 | 0 |
| rs143544421 | 9 | 136501599 | g | a | 5e-04 | 0.123 | 0.156 | 0.4303 | 1 | 0 |
| rs78445536 | 9 | 136501746 | g | a | 5e-04 | -0.0666 | 0.0572 | 0.2443 | 1 | 0 |
| rs146922432 | 9 | 136501768 | g | a | 3e-04 | -0.0174 | 0.144 | 0.9042 | 1 | 0 |
| rs76819676 | 9 | 136507375 | g | a | 2e-04 | -0.0232 | 0.111 | 0.8347 | 1 | 0 |
| rs200430427 | 9 | 136507456 | c | t | 4e-04 | -0.0581 | 0.154 | 0.706 | 1 | 0 |
| rs143535251 | 9 | 136507474 | c | t | 0.0013 | 0.0197 | 0.0248 | 0.4267 | 1 | 0 |
| rs200628504 | 9 | 136507528 | c | t | 4e-04 | 0.0516 | 0.128 | 0.6857 | 1 | 0 |
| rs5321 | 9 | 136507559 | g | c | 5e-04 | 0.0625 | 0.069 | 0.3652 | 1 | 0 |
| rs199734841 | 9 | 136507586 | g | c | 4e-04 | -0.176 | 0.2 | 0.3791 | 1 | 0 |
| rs13306301 | 9 | 136508640 | g | a | 1e-04 | -0.549 | 0.282 | 0.05178 | 1 | 0 |
| rs5324 | 9 | 136508658 | g | a | 6e-04 | -0.0219 | 0.0384 | 0.5682 | 1 | 0 |
| rs145655199 | 9 | 136508682 | g | a | 0.001 | -0.0173 | 0.18 | 0.9233 | 1 | 0 |
| rs201681337 | 9 | 136508691 | g | a | 0.002 | 0.0305 | 0.0754 | 0.6864 | 1 | 0 |
| rs75215331 | 9 | 136513028 | c | t | 0.0034 | 0.0146 | 0.0168 | 0.3834 | 1 | 0 |
| rs41316996 | 9 | 136521654 | g | a | 0.0032 | 0.019 | 0.0145 | 0.1918 | 1 | 0 |
| rs144040856 | 9 | 136521738 | g | a | 3e-04 | -0.0756 | 0.135 | 0.5758 | 1 | 0 |
| rs141021210 | 9 | 136521751 | a | g | 0 | -0.131 | 0.997 | 0.8956 | 1 | 0 |
| rs201973877 | 9 | 136522317 | t | c | 0.0003 | -0.0251 | 0.0565 | 0.6566 | 1 | 0 |
| rs75512464 | 9 | 136523487 | a | t | 0.002 | -0.0264 | 0.0236 | 0.2621 | 1 | 0 |
| rs76316834 | 9 | 136523555 | g | a | 4e-04 | 0.0484 | 0.13 | 0.709 | 1 | 0 |
| rs3025380 | 9 | 136501756 | g | c | 0.0045 | -0.0884 | 0.0119 | 1.096e-13 | 2 | -0.082 |
| rs74853476 | 9 | 136501834 | t | c | 0.0021 | -0.0774 | 0.0181 | 1.973e-05 | 2 | -0.082 |
| rs142383279 | 9 | 136507332 | g | a | 0.0018 | -0.0652 | 0.0194 | 0.0007811 | 2 | -0.082 |
| rs145059403 | 9 | 136507425 | g | a | 0.001 | -0.102 | 0.0261 | 9.144e-05 | 2 | -0.082 |
| rs148439785 | 9 | 136521726 | g | a | 8e-04 | -0.0595 | 0.0313 | 0.05706 | 2 | -0.082 |
| rs151228388 | 9 | 136522272 | a | g | 0.0013 | -0.317 | 0.102 | 0.001909 | 2 | -0.082 |

**Supplementary Table 15** Clustering results of rare variants within the NPR1 gene with SBP

| rsID | Chr | Position | Major allele | Minor allele | MAF | Effect | SE | P-value | Cluster membership | Cluster mean |
| --- | --- | --- | --- | --- | --- | --- | --- | --- | --- | --- |
| rs56019647 | 1 | 153652129 | c | t | 1e-04 | -0.113 | 0.165 | 0.4947 | 1 | 0 |
| rs28730726 | 1 | 153653757 | g | c | 0.0016 | -0.0175 | 0.0432 | 0.6842 | 1 | 0 |
| rs199612927 | 1 | 153654234 | g | a | 2e-04 | 0.0688 | 0.0871 | 0.4298 | 1 | 0 |
| rs140425746 | 1 | 153655966 | g | a | 2e-04 | -0.0333 | 0.154 | 0.8287 | 1 | 0 |
| rs201746049 | 1 | 153656216 | a | g | 0.0003 | -0.033 | 0.195 | 0.8658 | 1 | 0 |
| rs115938602 | 1 | 153656228 | c | a | 0 | -0.139 | 0.29 | 0.6323 | 1 | 0 |
| rs149202797 | 1 | 153658306 | c | a | 2e-04 | -0.0203 | 0.136 | 0.8808 | 1 | 0 |
| rs116775696 | 1 | 153659550 | a | g | 0 | -0.333 | 0.458 | 0.4672 | 1 | 0 |
| rs139174442 | 1 | 153660154 | c | g | 10.0e-05 | 0.297 | 0.35 | 0.3973 | 1 | 0 |
| rs201787421 | 1 | 153660625 | g | a | 2e-04 | -0.68 | 0.316 | 0.0312 | 1 | 0 |
| rs61757359 | 1 | 153658297 | g | a | 0.0034 | -0.0819 | 0.014 | 4.487e-09 | 2 | -0.0846 |
| rs61758562 | 1 | 153659131 | g | a | 5e-04 | -0.133 | 0.0495 | 0.007185 | 2 | -0.0846 |
| rs116245325 | 1 | 153665650 | c | t | 8e-04 | 0.166 | 0.0287 | 7.686e-09 | 3 | 0.164 |

**Supplementary Table 16** Clustering results of rare variants within the PLCB3 gene with SBP

| rsID | Chr | Position | Major allele | Minor allele | MAF | Effect | SE | P-value | Cluster membership | Cluster mean |
| --- | --- | --- | --- | --- | --- | --- | --- | --- | --- | --- |
| rs200996360 | 11 | 640253 | c | g | 0.0003 | -0.03 | 0.0527 | 0.5689 | 1 | 0 |
| rs111961110 | 11 | 64021930 | a | g | 0.0003 | -0.14 | 0.207 | 0.4974 | 1 | 0 |
| rs144191345 | 11 | 64022785 | c | t | 2e-04 | -0.0212 | 0.108 | 0.8442 | 1 | 0 |
| rs199645363 | 11 | 64022873 | c | t | 3e-04 | 0.0912 | 0.101 | 0.3668 | 1 | 0 |
| rs138400940 | 11 | 64022905 | c | t | 1e-04 | -0.381 | 0.576 | 0.5084 | 1 | 0 |
| rs188572550 | 11 | 64026357 | g | a | 0 | 0.801 | 0.704 | 0.2551 | 1 | 0 |
| rs201842672 | 11 | 64027567 | g | t | 0.0038 | -0.0627 | 0.0711 | 0.3777 | 1 | 0 |
| rs200923408 | 11 | 64028916 | g | a | 3e-04 | -0.538 | 0.235 | 0.02196 | 1 | 0 |
| rs200263631 | 11 | 64031571 | t | c | 0.0003 | 0.0847 | 0.0703 | 0.2283 | 1 | 0 |
| rs79573066 | 11 | 64032784 | g | a | 0.0036 | 0.0262 | 0.0696 | 0.7066 | 1 | 0 |
| rs61757725 | 11 | 64033360 | g | t | 0.009 | 0.0786 | 0.0599 | 0.1896 | 1 | 0 |
| rs148059922 | 11 | 64033391 | g | c | 4e-04 | -0.0149 | 0.131 | 0.9093 | 1 | 0 |
| rs201342752 | 11 | 64033986 | c | t | 0.002 | -0.0891 | 0.0706 | 0.2068 | 1 | 0 |
| rs141163685 | 11 | 64034734 | g | t | 0.0031 | 0.249 | 0.113 | 0.02777 | 1 | 0 |
| rs113554478 | 11 | 64034975 | g | a | 0.0011 | 0.00854 | 0.102 | 0.9333 | 1 | 0 |
| rs117874826 | 11 | 64027666 | a | c | 0.0138 | 0.0467 | 0.00729 | 1.535e-10 | 2 | 0.0529 |
| rs145502455 | 11 | 64031030 | g | a | 0.0054 | 0.0723 | 0.0118 | 8.563e-10 | 2 | 0.0529 |
| rs142330950 | 11 | 64032945 | g | t | 0.0021 | 0.0456 | 0.0182 | 0.01236 | 2 | 0.0529 |

**Supplementary Table 17** Clustering results of rare variants within the DBH gene with DBP

| rsID | Chr | Position | Major allele | Minor allele | MAF | Effect | SE | P-value | Cluster membership | Cluster mean |
| --- | --- | --- | --- | --- | --- | --- | --- | --- | --- | --- |
| rs76856960 | 9 | 136501569 | g | a | 0.0046 | -0.00876 | 0.0138 | 0.5248 | 1 | 0 |
| rs143544421 | 9 | 136501599 | g | a | 5e-04 | 0.0145 | 0.156 | 0.9262 | 1 | 0 |
| rs78445536 | 9 | 136501746 | g | a | 5e-04 | -0.0651 | 0.0572 | 0.255 | 1 | 0 |
| rs146922432 | 9 | 136501768 | g | a | 3e-04 | -0.125 | 0.144 | 0.3888 | 1 | 0 |
| rs76819676 | 9 | 136507375 | g | a | 2e-04 | 0.0357 | 0.111 | 0.7477 | 1 | 0 |
| rs200430427 | 9 | 136507456 | c | t | 4e-04 | 0.0385 | 0.154 | 0.8029 | 1 | 0 |
| rs143535251 | 9 | 136507474 | c | t | 0.0013 | 0.00881 | 0.0248 | 0.7226 | 1 | 0 |
| rs200628504 | 9 | 136507528 | c | t | 4e-04 | -0.081 | 0.128 | 0.5253 | 1 | 0 |
| rs5321 | 9 | 136507559 | g | c | 5e-04 | 0.0583 | 0.069 | 0.3984 | 1 | 0 |
| rs199734841 | 9 | 136507586 | g | c | 4e-04 | -0.225 | 0.2 | 0.2601 | 1 | 0 |
| rs13306301 | 9 | 136508640 | g | a | 1e-04 | -0.753 | 0.283 | 0.007722 | 1 | 0 |
| rs5324 | 9 | 136508658 | g | a | 6e-04 | -0.0431 | 0.0385 | 0.2625 | 1 | 0 |
| rs145655199 | 9 | 136508682 | g | a | 0.001 | 0.129 | 0.18 | 0.473 | 1 | 0 |
| rs201681337 | 9 | 136508691 | g | a | 0.002 | 0.0397 | 0.0755 | 0.5993 | 1 | 0 |
| rs75215331 | 9 | 136513028 | c | t | 0.0034 | 0.00505 | 0.0168 | 0.7633 | 1 | 0 |
| rs41316996 | 9 | 136521654 | g | a | 0.0032 | 0.00855 | 0.0146 | 0.557 | 1 | 0 |
| rs148439785 | 9 | 136521726 | g | a | 8e-04 | -0.0323 | 0.0313 | 0.3019 | 1 | 0 |
| rs144040856 | 9 | 136521738 | g | a | 3e-04 | -0.217 | 0.135 | 0.1089 | 1 | 0 |
| rs141021210 | 9 | 136521751 | a | g | 0 | -1.023 | 0.995 | 0.3037 | 1 | 0 |
| rs201973877 | 9 | 136522317 | t | c | 0.0003 | -0.0286 | 0.0566 | 0.6134 | 1 | 0 |
| rs75512464 | 9 | 136523487 | a | t | 0.002 | -0.025 | 0.0236 | 0.2893 | 1 | 0 |
| rs76316834 | 9 | 136523555 | g | a | 4e-04 | 0.015 | 0.13 | 0.9078 | 1 | 0 |
| rs3025380 | 9 | 136501756 | g | c | 0.0045 | -0.103 | 0.0119 | 4.287e-18 | 2 | -0.0927 |
| rs74853476 | 9 | 136501834 | t | c | 0.0021 | -0.0954 | 0.0182 | 1.529e-07 | 2 | -0.0927 |
| rs142383279 | 9 | 136507332 | g | a | 0.0018 | -0.0701 | 0.0194 | 0.0003159 | 2 | -0.0927 |
| rs145059403 | 9 | 136507425 | g | a | 0.001 | -0.0829 | 0.0262 | 0.001551 | 2 | -0.0927 |
| rs151228388 | 9 | 136522272 | a | g | 0.0013 | -0.226 | 0.102 | 0.02711 | 2 | -0.0927 |

**Supplementary Table 18** Clustering results of rare variants within the PLCB3 gene with DBP

| rsID | Chr | Position | Major allele | Minor allele | MAF | Effect | SE | P-value | Cluster membership | Cluster mean |
| --- | --- | --- | --- | --- | --- | --- | --- | --- | --- | --- |
| rs200996360 | 11 | 640253 | c | g | 0.0003 | 0.0237 | 0.0528 | 0.653 | 1 | 0 |
| rs111961110 | 11 | 64021930 | a | g | 0.0003 | 0.00639 | 0.207 | 0.9754 | 1 | 0 |
| rs144191345 | 11 | 64022785 | c | t | 2e-04 | 0.0759 | 0.108 | 0.4822 | 1 | 0 |
| rs199645363 | 11 | 64022873 | c | t | 3e-04 | 0.0863 | 0.101 | 0.3935 | 1 | 0 |
| rs138400940 | 11 | 64022905 | c | t | 1e-04 | -0.858 | 0.576 | 0.1362 | 1 | 0 |
| rs188572550 | 11 | 64026357 | g | a | 0 | 0.624 | 0.704 | 0.3759 | 1 | 0 |
| rs201842672 | 11 | 64027567 | g | t | 0.0038 | -0.0781 | 0.0711 | 0.2721 | 1 | 0 |
| rs200923408 | 11 | 64028916 | g | a | 3e-04 | -0.194 | 0.235 | 0.4077 | 1 | 0 |
| rs79573066 | 11 | 64032784 | g | a | 0.0036 | 0.0198 | 0.0697 | 0.7768 | 1 | 0 |
| rs61757725 | 11 | 64033360 | g | t | 0.009 | 0.0537 | 0.06 | 0.3709 | 1 | 0 |
| rs148059922 | 11 | 64033391 | g | c | 4e-04 | -0.154 | 0.131 | 0.2409 | 1 | 0 |
| rs201342752 | 11 | 64033986 | c | t | 0.002 | -0.0181 | 0.0706 | 0.7979 | 1 | 0 |
| rs141163685 | 11 | 64034734 | g | t | 0.0031 | 0.214 | 0.113 | 0.05838 | 1 | 0 |
| rs113554478 | 11 | 64034975 | g | a | 0.0011 | 0.0951 | 0.102 | 0.3518 | 1 | 0 |
| rs117874826 | 11 | 64027666 | a | c | 0.0138 | 0.041 | 0.00731 | 2.069e-08 | 2 | 0.0473 |
| rs145502455 | 11 | 64031030 | g | a | 0.0054 | 0.0668 | 0.0118 | 1.619e-08 | 2 | 0.0473 |
| rs200263631 | 11 | 64031571 | t | c | 0.0003 | 0.121 | 0.0704 | 0.08613 | 2 | 0.0473 |
| rs142330950 | 11 | 64032945 | g | t | 0.0021 | 0.0367 | 0.0182 | 0.04445 | 2 | 0.0473 |

**Supplementary Table 19** Clustering results of rare variants within the CEP120 gene with PP

| rsID | Chr | Position | Major allele | Minor allele | MAF | Effect | SE | P-value | Cluster membership | Cluster mean |
| --- | --- | --- | --- | --- | --- | --- | --- | --- | --- | --- |
| rs145436175 | 5 | 122682248 | t | c | 0.0008 | 0.233 | 0.268 | 0.385 | 1 | 0 |
| rs140306974 | 5 | 122685717 | g | a | 0.0011 | 0.0594 | 0.126 | 0.6367 | 1 | 0 |
| rs200061679 | 5 | 122685731 | t | a | 0.0015 | 0.0692 | 0.0388 | 0.07491 | 1 | 0 |
| rs142792779 | 5 | 122708381 | c | t | 3e-04 | -0.0136 | 0.213 | 0.9492 | 1 | 0 |
| rs139865050 | 5 | 122713092 | c | g | 0 | -0.192 | 0.707 | 0.7861 | 1 | 0 |
| rs74938108 | 5 | 122713159 | c | t | 0.0016 | -0.0257 | 0.0905 | 0.7765 | 1 | 0 |
| rs201600892 | 5 | 122713191 | c | g | 10.0e-05 | 0.0767 | 0.447 | 0.8637 | 1 | 0 |
| rs61744334 | 5 | 122714044 | t | c | 0.0006 | -0.0201 | 0.0684 | 0.7689 | 1 | 0 |
| rs144490830 | 5 | 122714104 | g | c | 2e-04 | 0.37 | 0.179 | 0.03908 | 1 | 0 |
| rs147277049 | 5 | 122720724 | t | c | 0.0006 | -0.0181 | 0.036 | 0.6152 | 1 | 0 |
| rs200450605 | 5 | 122725693 | g | a | 0.0017 | 0.016 | 0.0229 | 0.4841 | 1 | 0 |
| rs201571160 | 5 | 122725754 | c | g | 0.0003 | 0.166 | 0.316 | 0.6004 | 1 | 0 |
| rs114281792 | 5 | 122725761 | t | c | 0.0067 | -0.00233 | 0.0098 | 0.8124 | 1 | 0 |
| rs61747983 | 5 | 122725768 | g | a | 1e-04 | 0.0156 | 0.112 | 0.8895 | 1 | 0 |
| rs201955087 | 5 | 122727015 | c | t | 0.0013 | 0.274 | 0.106 | 0.009927 | 1 | 0 |
| rs147273517 | 5 | 122748194 | t | c | 10.0e-05 | 0.36 | 0.162 | 0.02599 | 1 | 0 |
| rs202103949 | 5 | 122748198 | t | c | 0.0004 | -0.029 | 0.121 | 0.8106 | 1 | 0 |
| rs199793672 | 5 | 122758665 | t | c | 10.0e-05 | -0.479 | 0.503 | 0.3404 | 1 | 0 |
| rs2303720 | 5 | 122682334 | c | t | 0.0291 | -0.0418 | 0.00482 | 4.532e-18 | 2 | -0.0435 |
| rs114280473 | 5 | 122714092 | g | a | 0.0063 | -0.0632 | 0.011 | 8.076e-09 | 2 | -0.0435 |
| rs189429890 | 5 | 122729025 | c | t | 0.0065 | -0.0373 | 0.00999 | 0.0001886 | 2 | -0.0435 |

**Supplementary Table 20** Clustering results of rare variants within COL21A1 gene with PP

| rsID | Chr | Position | Major allele | Minor allele | MAF | Effect | SE | P-value | Cluster membership | Cluster mean |
| --- | --- | --- | --- | --- | --- | --- | --- | --- | --- | --- |
| rs200478915 | 6 | 55923968 | g | c | 8e-04 | -0.0449 | 0.0323 | 0.1652 | 1 | 0 |
| rs200564236 | 6 | 55925008 | g | a | 8e-04 | -0.0212 | 0.162 | 0.8957 | 1 | 0 |
| rs199722485 | 6 | 55925588 | t | g | 0.0005 | -0.0128 | 0.106 | 0.9038 | 1 | 0 |
| rs200674177 | 6 | 55925689 | g | a | 2e-04 | -0.0981 | 0.189 | 0.6046 | 1 | 0 |
| rs201839603 | 6 | 55925762 | a | c | 10.0e-05 | 0.0193 | 0.577 | 0.9733 | 1 | 0 |
| rs9464337 | 6 | 55925801 | g | t | 0.0054 | 0.000204 | 0.0116 | 0.986 | 1 | 0 |
| rs201892311 | 6 | 55926464 | g | c | 1e-04 | 0.426 | 0.707 | 0.547 | 1 | 0 |
| rs199910287 | 6 | 55935556 | g | a | 6e-04 | 0.054 | 0.0445 | 0.2255 | 1 | 0 |
| rs191626317 | 6 | 55939059 | g | t | 0.002 | 0.0424 | 0.102 | 0.6783 | 1 | 0 |
| rs75605879 | 6 | 55966311 | a | g | 10.0e-05 | -0.147 | 0.107 | 0.169 | 1 | 0 |
| rs202115077 | 6 | 55988871 | g | t | 0.002 | 0.00866 | 0.0186 | 0.6409 | 1 | 0 |
| rs201267383 | 6 | 56006611 | t | c | 0.0003 | -0.077 | 0.0817 | 0.3461 | 1 | 0 |
| rs35583895 | 6 | 56006732 | a | g | 0.0081 | 0.00138 | 0.0479 | 0.9771 | 1 | 0 |
| rs200361985 | 6 | 56029264 | g | a | 0.002 | 0.0616 | 0.0546 | 0.2593 | 1 | 0 |
| rs142653960 | 6 | 56035494 | c | t | 4e-04 | -0.0656 | 0.0525 | 0.2115 | 1 | 0 |
| rs199532612 | 6 | 56035853 | a | t | 0.0017 | -0.0186 | 0.0217 | 0.3925 | 1 | 0 |
| rs200708113 | 6 | 56035881 | c | t | 5e-04 | -0.0112 | 0.0509 | 0.8259 | 1 | 0 |
| rs202026963 | 6 | 56035909 | g | a | 1e-04 | 0.0193 | 0.242 | 0.9364 | 1 | 0 |
| rs147394600 | 6 | 56047400 | g | a | 1e-04 | -0.159 | 0.164 | 0.331 | 1 | 0 |
| rs200999181 | 6 | 55935568 | c | a | 0.0012 | 0.336 | 0.0244 | 3.471e-43 | 2 | 0.335 |
| rs115079907 | 6 | 55924005 | c | t | 0.0015 | 0.207 | 0.0248 | 5.573e-17 | 3 | 0.171 |
| rs76146749 | 6 | 55925783 | t | a | 7e-04 | 0.198 | 0.0782 | 0.01144 | 3 | 0.171 |
| rs200401514 | 6 | 55989091 | c | t | 4e-04 | 0.124 | 0.0552 | 0.02532 | 3 | 0.171 |
| rs2764043 | 6 | 56035643 | a | g | 0.0016 | 0.152 | 0.0203 | 7.82e-14 | 3 | 0.171 |

**Supplementary Table 21** Clustering results of rare variants within the NOX4 gene with PP

| rsID | Chr | Position | Major allele | Minor allele | MAF | Effect | SE | P-value | Cluster membership | Cluster mean |
| --- | --- | --- | --- | --- | --- | --- | --- | --- | --- | --- |
| rs115031759 | 11 | 89073269 | g | a | 1e-04 | 0.192 | 0.161 | 0.2323 | 1 | 0 |
| rs201165492 | 11 | 89106599 | c | t | 1e-04 | -0.274 | 0.576 | 0.6345 | 1 | 0 |
| rs149515506 | 11 | 89135492 | a | g | 10.0e-05 | 0.428 | 0.706 | 0.5445 | 1 | 0 |
| rs147350656 | 11 | 89177310 | c | t | 1e-04 | -0.184 | 0.236 | 0.4375 | 1 | 0 |
| rs142433357 | 11 | 89182609 | t | c | 10.0e-05 | -1.231 | 0.707 | 0.08181 | 1 | 0 |
| rs55977241 | 11 | 89182652 | c | t | 0.002 | 0.000803 | 0.109 | 0.9941 | 1 | 0 |
| rs145686545 | 11 | 89224387 | c | t | 2e-04 | 0.162 | 0.202 | 0.4229 | 1 | 0 |
| rs144215891 | 11 | 89069094 | t | c | 0.0014 | -0.0716 | 0.0302 | 0.01798 | 2 | -0.0962 |
| rs139341533 | 11 | 89182666 | c | a | 0.0043 | -0.0905 | 0.0126 | 8.085e-13 | 2 | -0.0962 |
| rs56061986 | 11 | 89182686 | t | c | 0.0029 | -0.113 | 0.0161 | 2.385e-12 | 2 | -0.0962 |

**Supplementary Table 22** Clustering results of rare variants within the DBH gene with HTN

| rsID | Chr | Position | Major allele | Minor allele | MAF | Z value | P-value | Cluster membership | Cluster mean |
| --- | --- | --- | --- | --- | --- | --- | --- | --- | --- |
| rs76856960 | 9 | 136501569 | g | a | 0.0035 | -0.511 | 0.6096 | 1 | 0 |
| rs143544421 | 9 | 136501599 | g | a | 2e-04 | -0.526 | 0.5989 | 1 | 0 |
| rs78445536 | 9 | 136501746 | g | a | 3e-04 | 0.234 | 0.815 | 1 | 0 |
| rs146922432 | 9 | 136501768 | g | a | 1e-04 | 1.257 | 0.2089 | 1 | 0 |
| rs76819676 | 9 | 136507375 | g | a | 1e-04 | -0.787 | 0.4313 | 1 | 0 |
| rs143535251 | 9 | 136507474 | c | t | 0.0014 | 0.397 | 0.6916 | 1 | 0 |
| rs200628504 | 9 | 136507528 | c | t | 2e-04 | -0.0675 | 0.9462 | 1 | 0 |
| rs5321 | 9 | 136507559 | g | c | 2e-04 | 0.226 | 0.8212 | 1 | 0 |
| rs199734841 | 9 | 136507586 | g | c | 2e-04 | 0.897 | 0.3697 | 1 | 0 |
| rs13306301 | 9 | 136508640 | g | a | 0 | -1.378 | 0.1683 | 1 | 0 |
| rs5324 | 9 | 136508658 | g | a | 8e-04 | -1.404 | 0.1604 | 1 | 0 |
| rs145655199 | 9 | 136508682 | g | a | 3e-04 | -1.247 | 0.2125 | 1 | 0 |
| rs201681337 | 9 | 136508691 | g | a | 2e-04 | 1.109 | 0.2676 | 1 | 0 |
| rs75215331 | 9 | 136513028 | c | t | 0.0025 | -0.0818 | 0.9348 | 1 | 0 |
| rs41316996 | 9 | 136521654 | g | a | 0.003 | 2.028 | 0.04261 | 1 | 0 |
| rs144040856 | 9 | 136521738 | g | a | 1e-04 | 0.0598 | 0.9523 | 1 | 0 |
| rs141021210 | 9 | 136521751 | a | g | 0 | -0.665 | 0.5059 | 1 | 0 |
| rs201973877 | 9 | 136522317 | t | c | 0.0002 | -0.923 | 0.356 | 1 | 0 |
| rs75512464 | 9 | 136523487 | a | t | 0.0014 | -0.289 | 0.7723 | 1 | 0 |
| rs148806316 | 9 | 136523534 | c | t | 0.0025 | 0.295 | 0.7679 | 1 | 0 |
| rs76316834 | 9 | 136523555 | g | a | 2e-04 | -0.39 | 0.6962 | 1 | 0 |
| rs77273740 | 9 | 136501728 | c | t | 0.0042 | -2.933 | 0.003353 | 2 | -2.612 |
| rs3025380 | 9 | 136501756 | g | c | 0.0045 | -5.297 | 1.176e-07 | 2 | -2.612 |
| rs74853476 | 9 | 136501834 | t | c | 0.0022 | -2.186 | 0.02885 | 2 | -2.612 |
| rs142383279 | 9 | 136507332 | g | a | 0.002 | -1.826 | 0.06781 | 2 | -2.612 |
| rs145059403 | 9 | 136507425 | g | a | 0.0012 | -3.468 | 0.0005248 | 2 | -2.612 |
| rs200430427 | 9 | 136507456 | c | t | 1e-04 | -2.306 | 0.02112 | 2 | -2.612 |
| rs148439785 | 9 | 136521726 | g | a | 0.0019 | -2.196 | 0.02806 | 2 | -2.612 |
| rs151228388 | 9 | 136522272 | a | g | 0.0003 | -2.399 | 0.01646 | 2 | -2.612 |

**Supplementary Table 23** Clustering results of rare variants within the NPR1 gene with HTN

| rsID | Chr | Position | Major allele | Minor allele | MAF | Z value | P-value | Cluster membership | Cluster mean |
| --- | --- | --- | --- | --- | --- | --- | --- | --- | --- |
| rs56019647 | 1 | 153652129 | c | t | 0 | -0.727 | 0.4675 | 1 | 0 |
| rs199612927 | 1 | 153654234 | g | a | 1e-04 | 0.468 | 0.6397 | 1 | 0 |
| rs201746049 | 1 | 153656216 | a | g | 10.0e-05 | -1.254 | 0.21 | 1 | 0 |
| rs115938602 | 1 | 153656228 | c | a | 0 | -1.174 | 0.2405 | 1 | 0 |
| rs149202797 | 1 | 153658306 | c | a | 1e-04 | 0.23 | 0.8182 | 1 | 0 |
| rs116775696 | 1 | 153659550 | a | g | 0 | 0.641 | 0.5213 | 1 | 0 |
| rs139174442 | 1 | 153660154 | c | g | 0 | 0.0493 | 0.9606 | 1 | 0 |
| rs201787421 | 1 | 153660625 | g | a | 1e-04 | -0.287 | 0.7739 | 1 | 0 |
| rs140425746 | 1 | 153655966 | g | a | 1e-04 | -1.607 | 0.1081 | 2 | -2.053 |
| rs61757359 | 1 | 153658297 | g | a | 0.0036 | -3.719 | 2e-04 | 2 | -2.053 |
| rs61758562 | 1 | 153659131 | g | a | 0.0011 | -1.693 | 0.09054 | 2 | -2.053 |
| rs116245325 | 1 | 153665650 | c | t | 8e-04 | 4.335 | 1.457e-05 | 3 | 4.334 |

**Supplementary Table 24** Clustering results of rare variants within the PLCB3 gene with HTN

| rsID | Chr | Position | Major allele | Minor allele | MAF | Z value | P-value | Cluster membership | Cluster mean |
| --- | --- | --- | --- | --- | --- | --- | --- | --- | --- |
| rs200996360 | 11 | 640253 | c | g | 0.0003 | -0.905 | 0.3653 | 1 | 0 |
| rs111961110 | 11 | 64021930 | a | g | 10.0e-05 | -0.917 | 0.3591 | 1 | 0 |
| rs144191345 | 11 | 64022785 | c | t | 1e-04 | 0.259 | 0.7958 | 1 | 0 |
| rs199645363 | 11 | 64022873 | c | t | 2e-04 | -0.587 | 0.5575 | 1 | 0 |
| rs138400940 | 11 | 64022905 | c | t | 1e-04 | -1.28 | 0.2004 | 1 | 0 |
| rs188572550 | 11 | 64026357 | g | a | 0 | -0.324 | 0.7457 | 1 | 0 |
| rs201842672 | 11 | 64027567 | g | t | 9e-04 | -0.58 | 0.5616 | 1 | 0 |
| rs200923408 | 11 | 64028916 | g | a | 1e-04 | -0.496 | 0.6198 | 1 | 0 |
| rs200263631 | 11 | 64031571 | t | c | 0.0002 | 1.033 | 0.3018 | 1 | 0 |
| rs79573066 | 11 | 64032784 | g | a | 2e-04 | 0.665 | 0.5063 | 1 | 0 |
| rs142330950 | 11 | 64032945 | g | t | 0.0021 | 2.175 | 0.02966 | 1 | 0 |
| rs61757725 | 11 | 64033360 | g | t | 3e-04 | 0.427 | 0.6694 | 1 | 0 |
| rs148059922 | 11 | 64033391 | g | c | 1e-04 | 1.308 | 0.1908 | 1 | 0 |
| rs201342752 | 11 | 64033986 | c | t | 5e-04 | 1.223 | 0.2215 | 1 | 0 |
| rs141163685 | 11 | 64034734 | g | t | 4e-04 | 0.867 | 0.3859 | 1 | 0 |
| rs113554478 | 11 | 64034975 | g | a | 1e-04 | 0.708 | 0.4787 | 1 | 0 |
| rs117874826 | 11 | 64027666 | a | c | 0.0308 | 5.712 | 1.119e-08 | 2 | 4.808 |
| rs145502455 | 11 | 64031030 | g | a | 0.0047 | 4.015 | 5.952e-05 | 2 | 4.808 |

**Supplementary Figures**

**Supplementary Figure 1** Heatmap of the average correlation matrix of the 60 simulated rare variants with the presence of LD structure

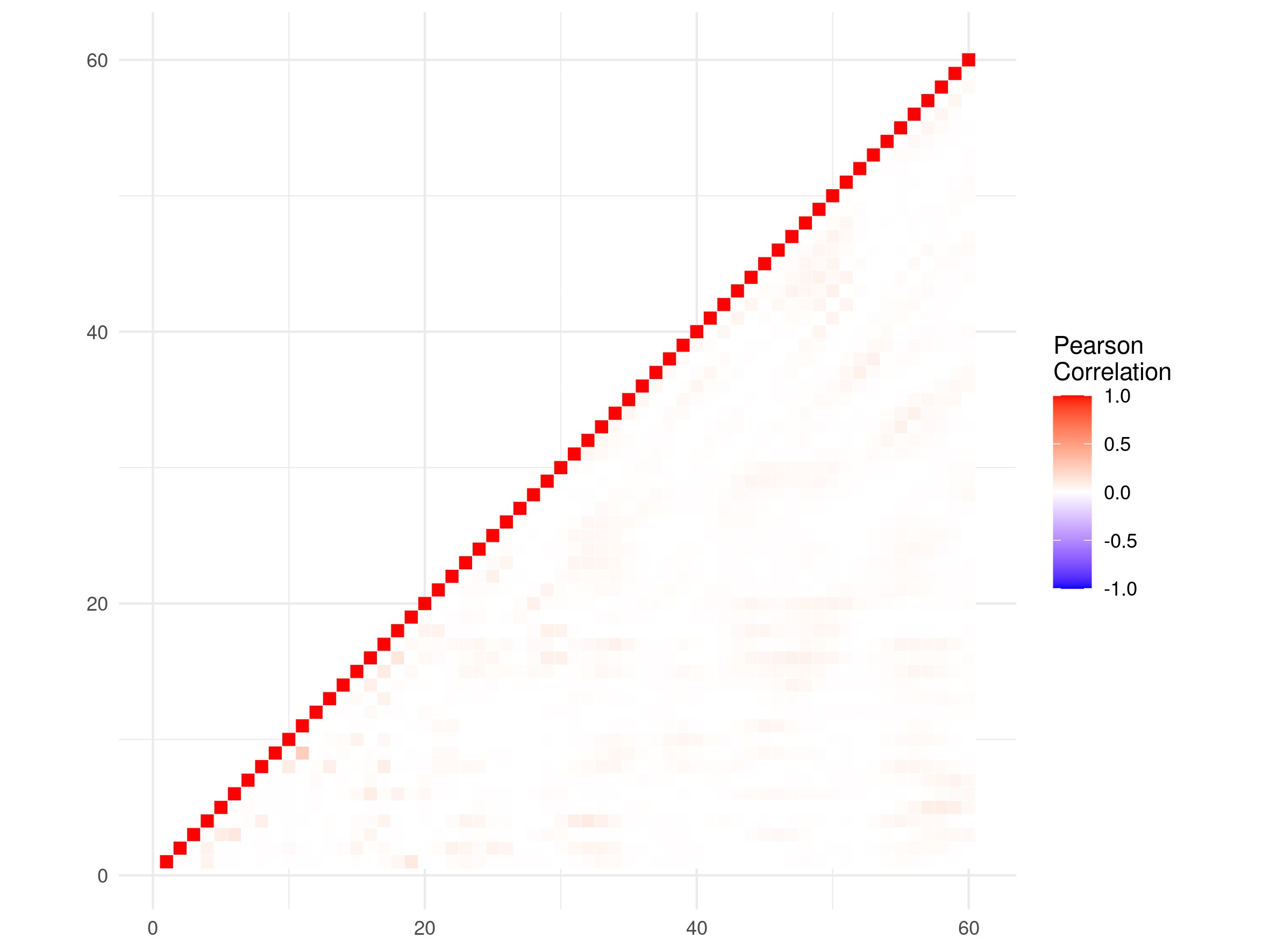

**Supplementary Figure 2** Boxplots of ARI values of simulation scenarios 1, 2, and 3 with combined weighting scheme with/without LD for a binary trait

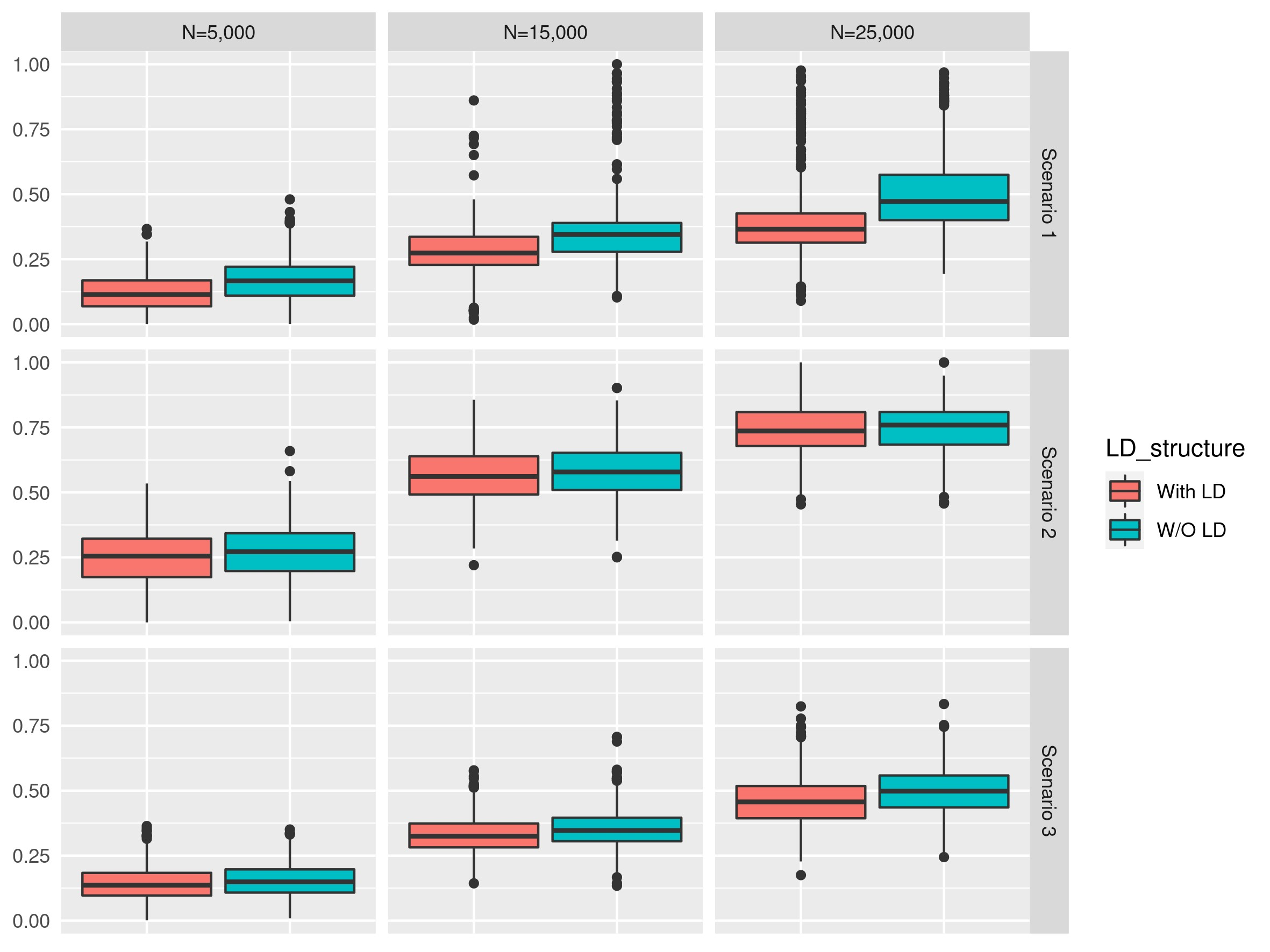

**Supplementary Figure 3** Boxplots of ARI values of simulation scenarios 4, 5, and 6 with combined weighting scheme with/without LD for a binary trait

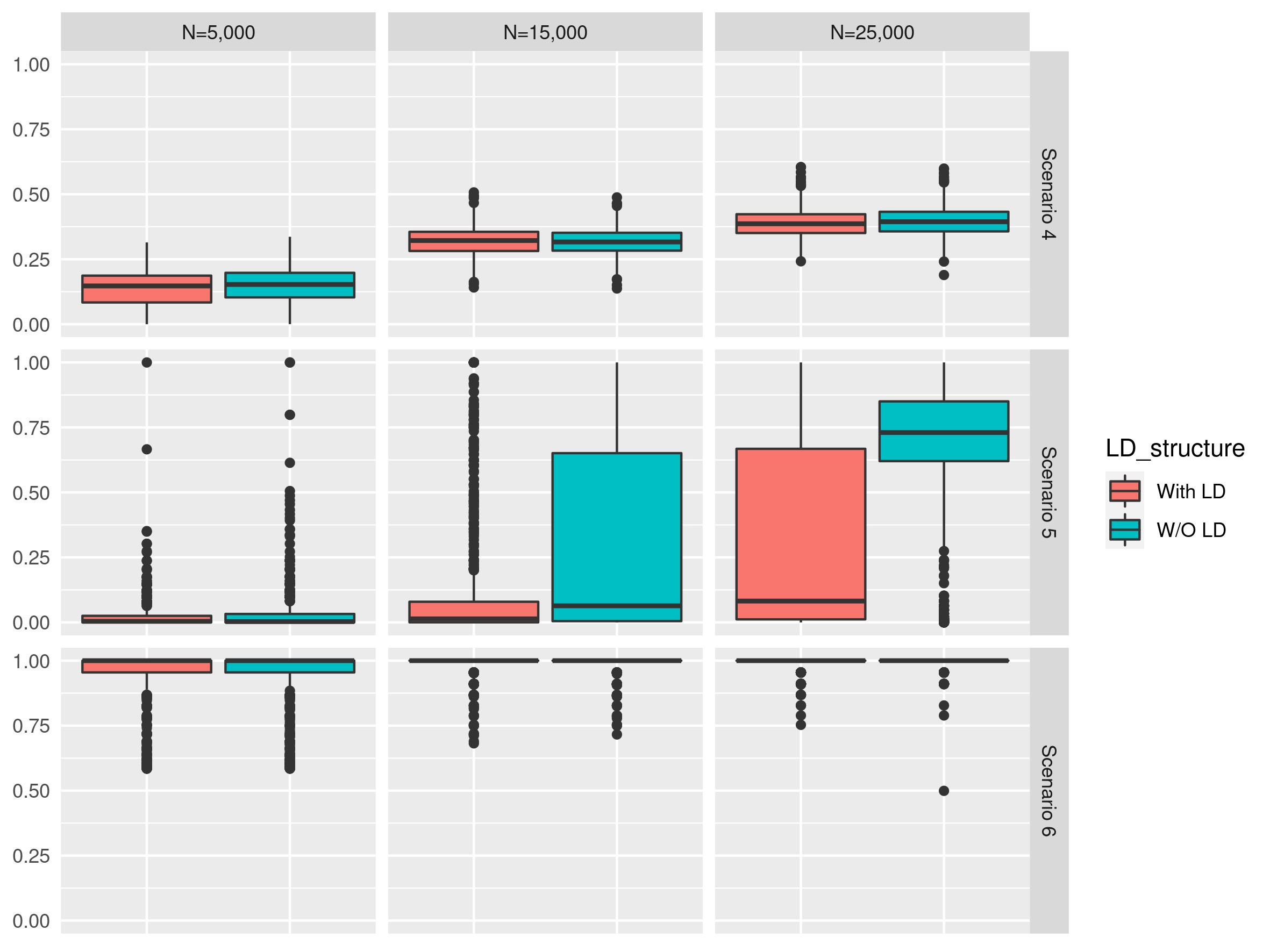

**Supplementary Figure 4** Boxplots of ARI values of simulation scenarios 1, 2, and 3 with equal, inverse SE and combined weighting schemes with the absence of LD for a continuous trait

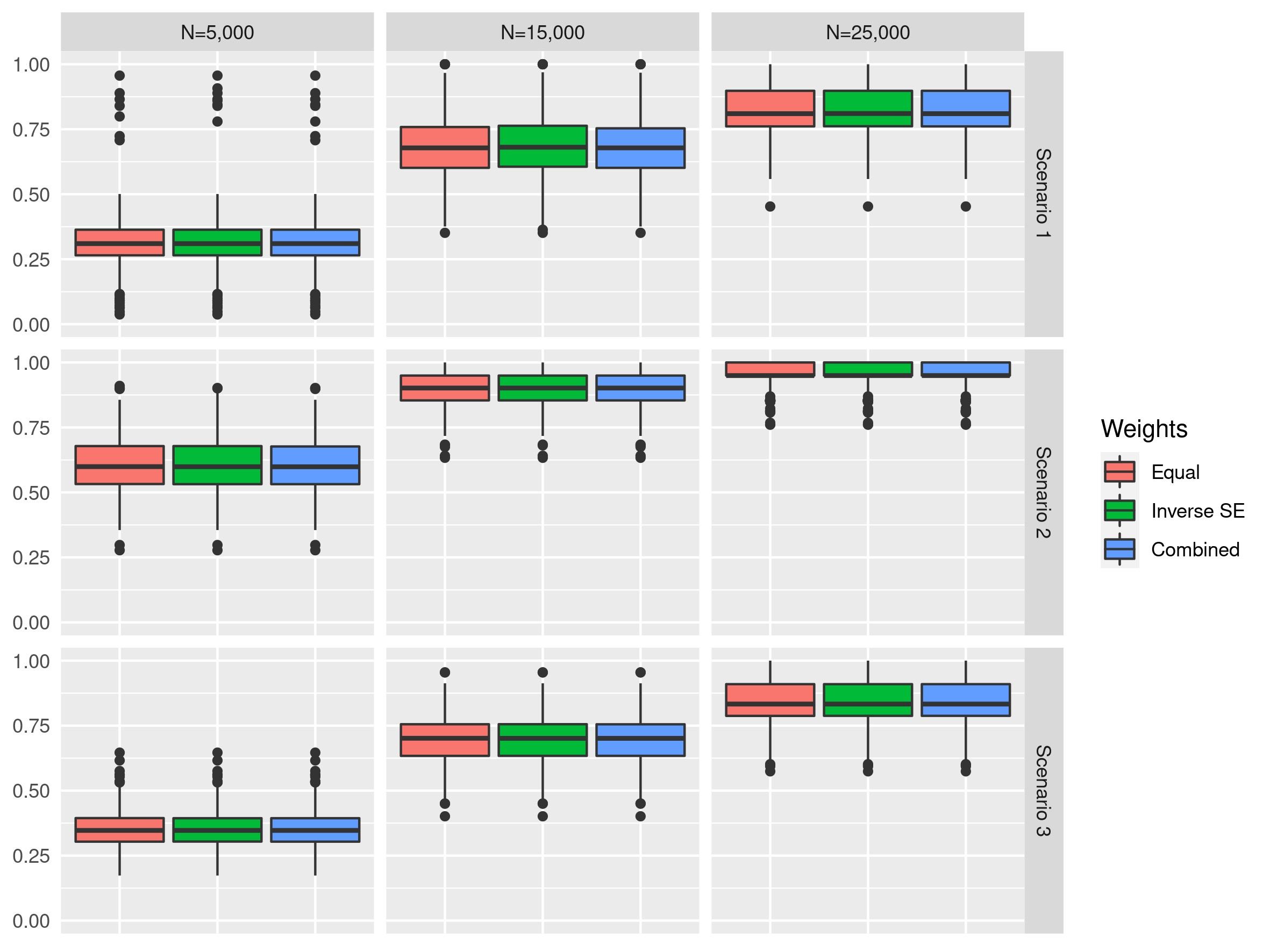

**Supplementary Figure 5** Boxplots of ARI values of simulation scenarios 4, 5, and 6 with equal, inverse SE and combined weighting schemes with the absence of LD for a continuous trait

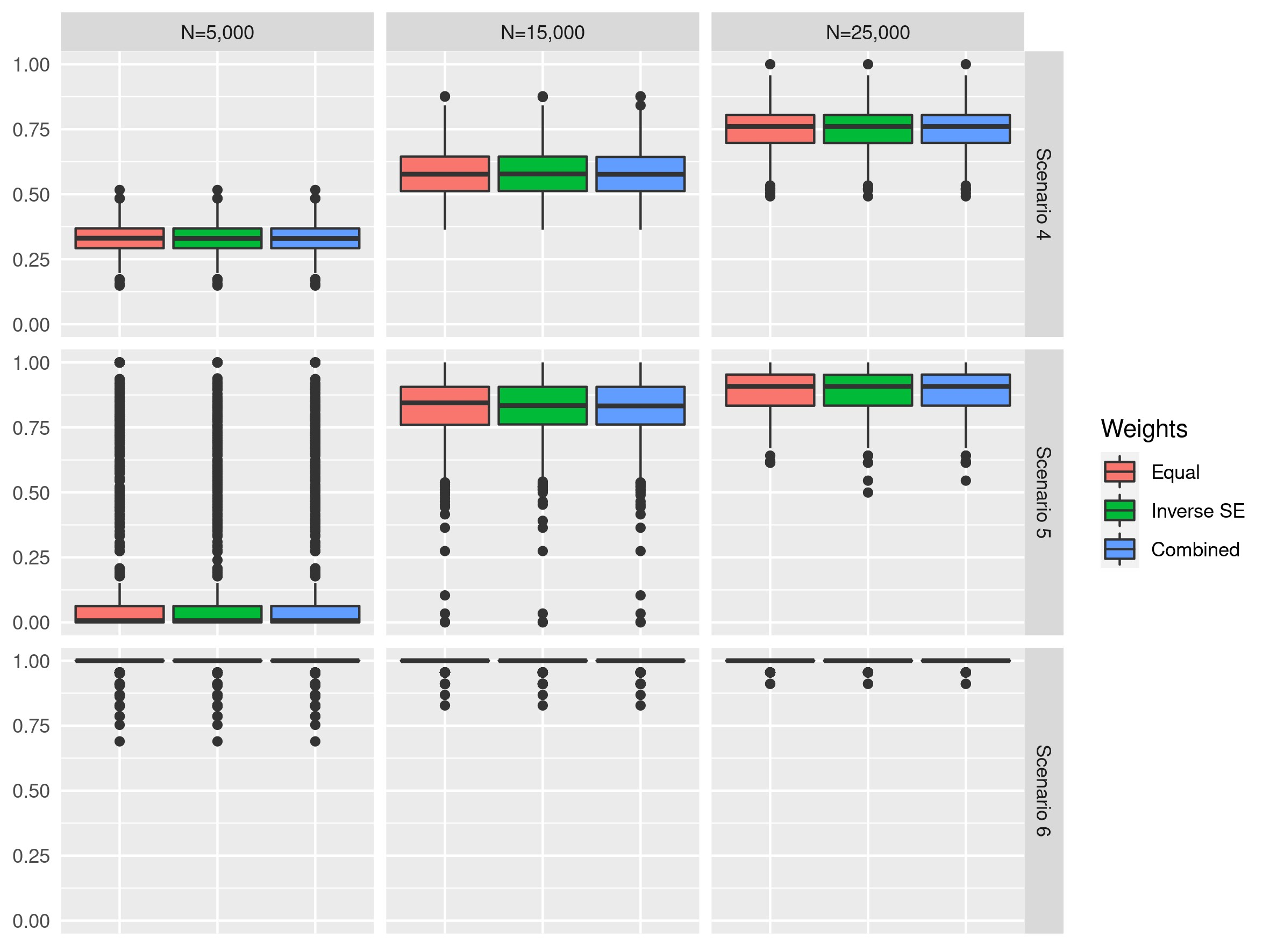

**Supplementary Figure 6** Boxplots of ARI values of simulation scenarios 1, 2, and 3 with equal, inverse SE and combined weighting schemes with the absence of LD for a binary trait

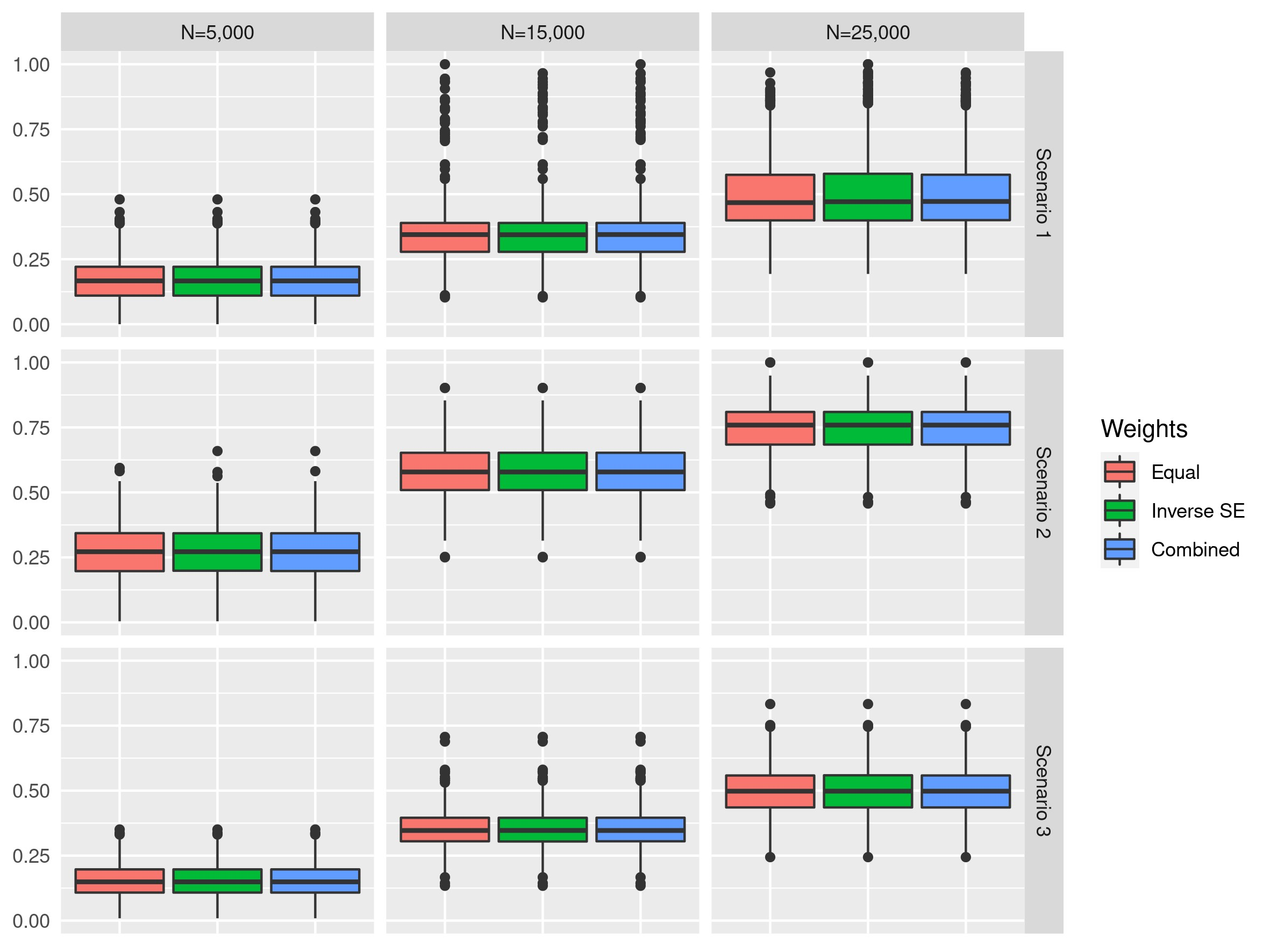

**Supplementary Figure 7** Boxplots of ARI values of simulation scenarios 4, 5, and 6 with equal, inverse SE and combined weighting schemes with the absence of LD for a binary trait

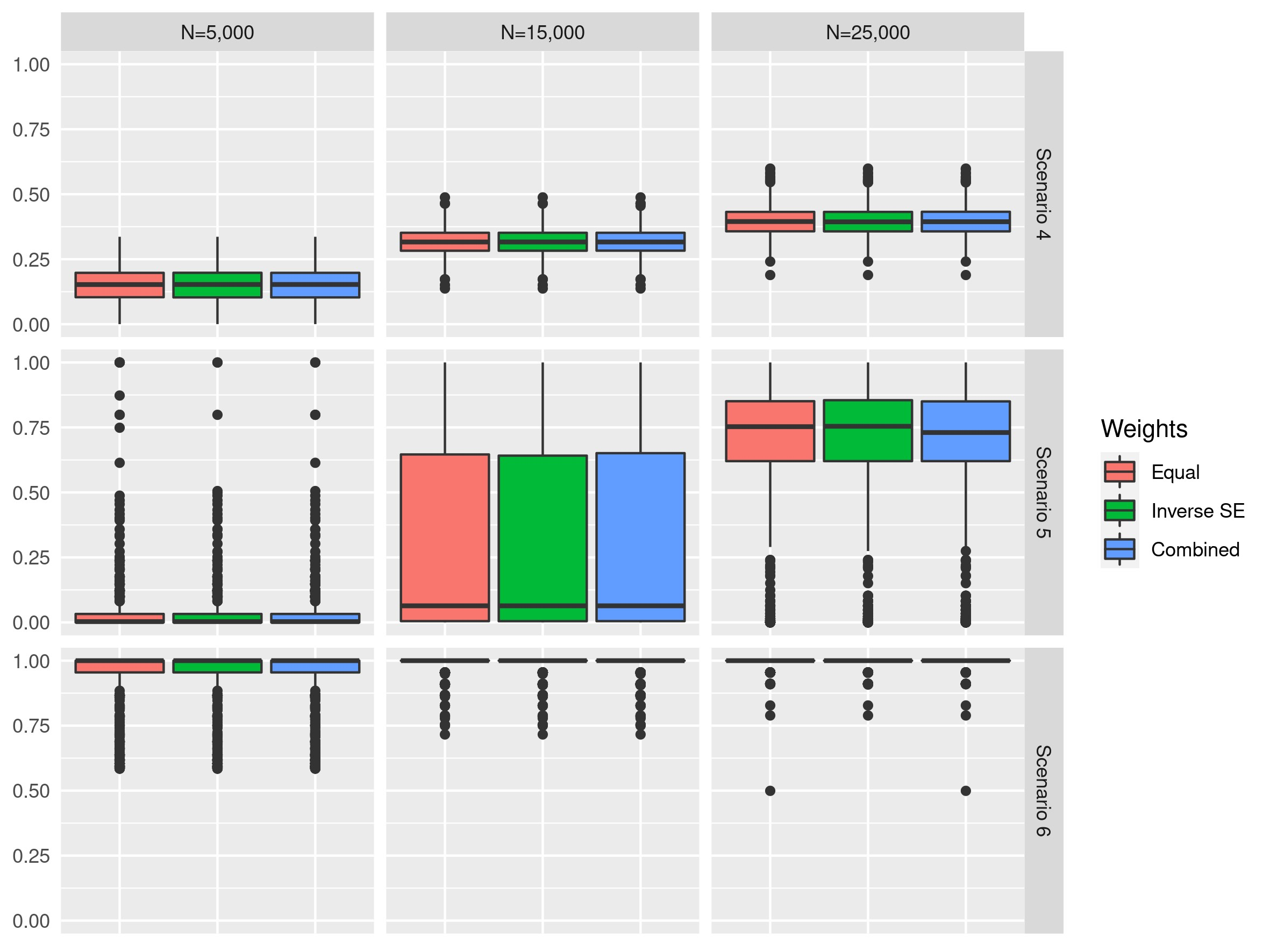

**Supplementary Figure 8** Boxplots of ARI values of simulation scenarios 1, 2, and 3 with equal, inverse SE and combined weighting schemes with the presence of LD for a continuous trait

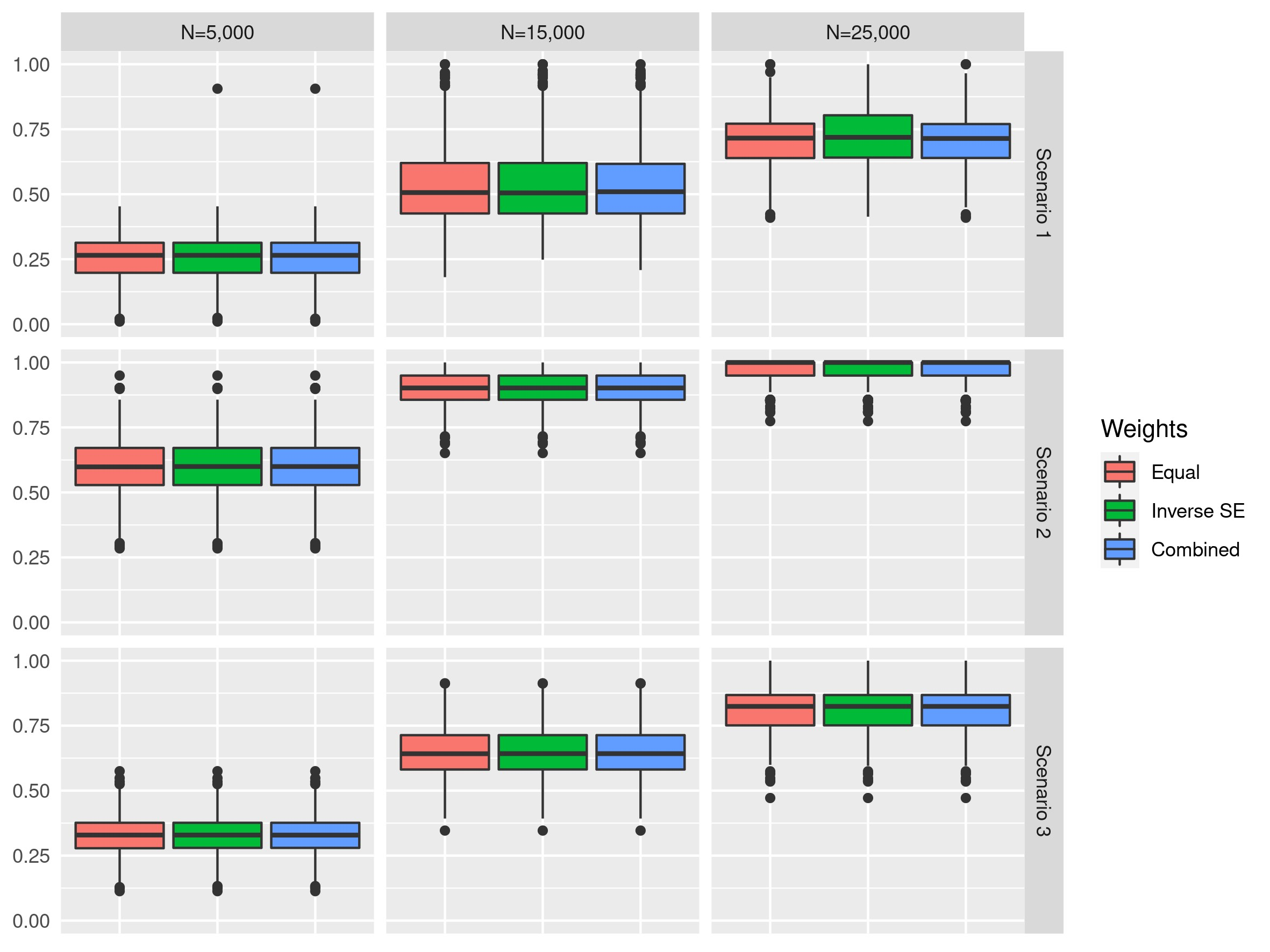

**Supplementary Figure 9** Boxplots of ARI values of simulation scenarios 4, 5, and 6 with equal, inverse SE and combined weighting schemes with the presence of LD for a continuous trait

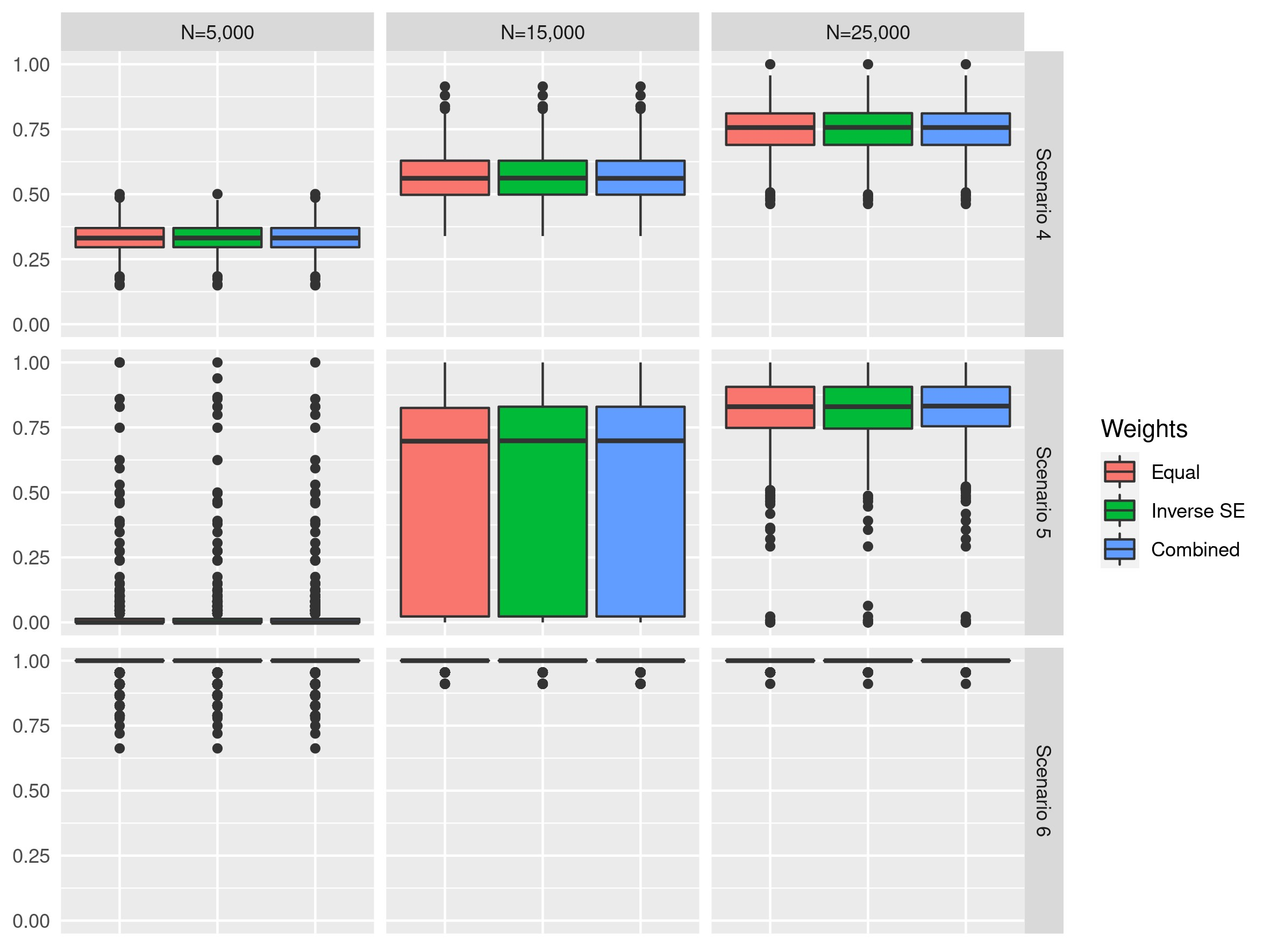

**Supplementary Figure 10** Boxplots of ARI values of simulation scenarios 1, 2, and 3 with equal, inverse SE and combined weighting schemes with the presence of LD for a binary trait

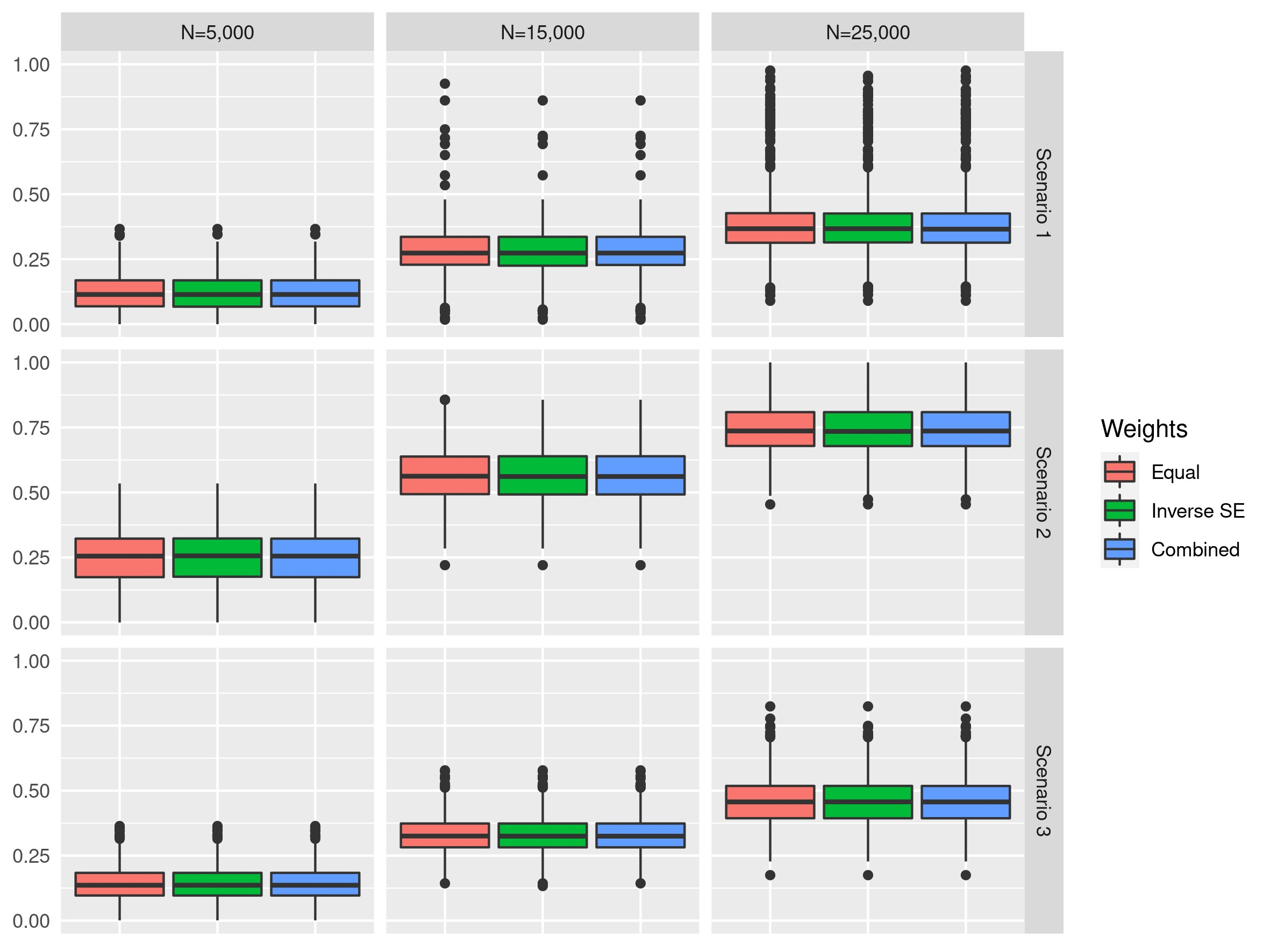

**Supplementary Figure 11** Boxplots of ARI values of simulation scenarios 4, 5, and 6 with equal, inverse SE and combined weighting schemes with the presence of LD for a binary trait

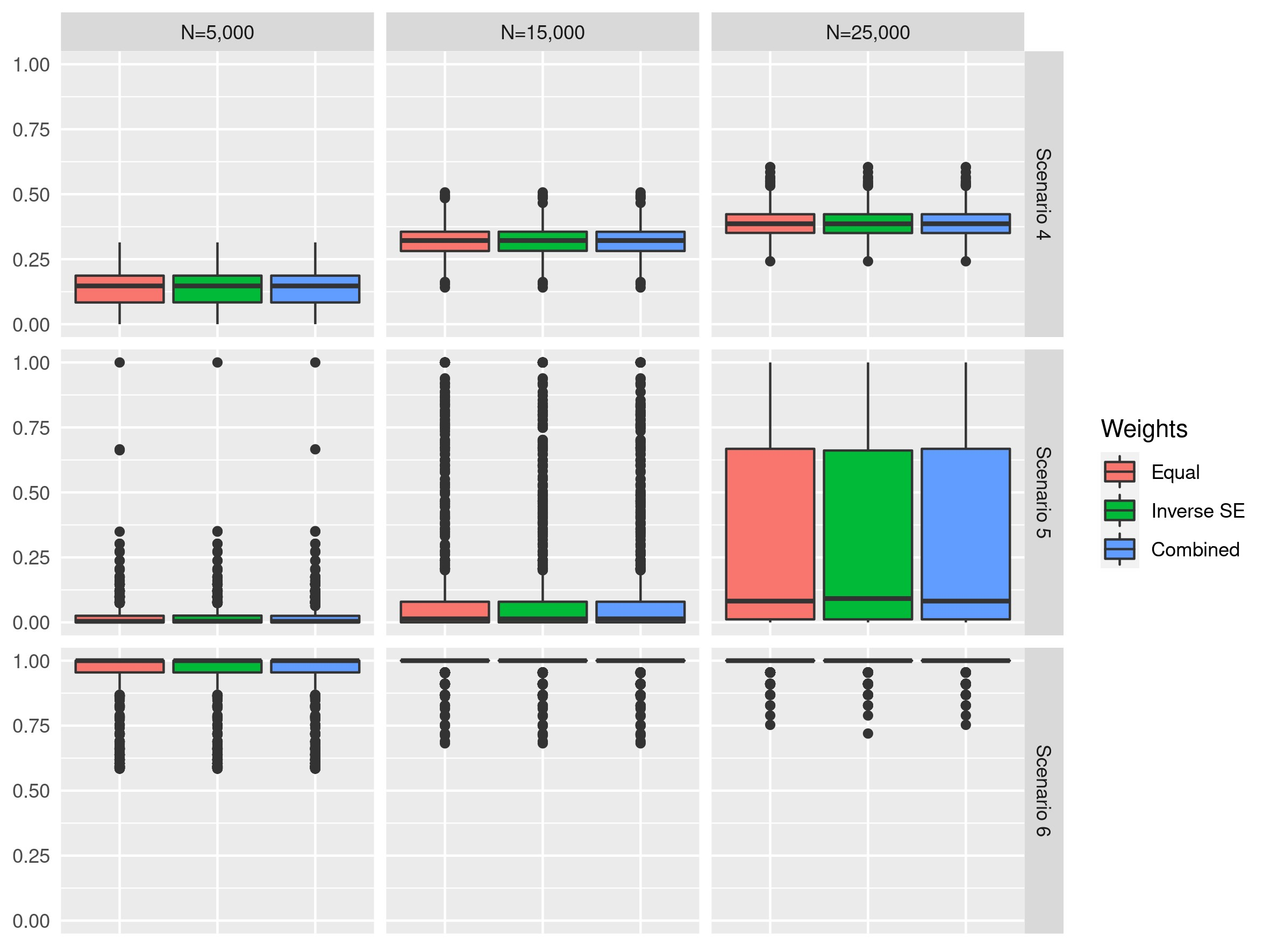
